# Quantifying the association between neutralising antibodies and protection from RSV disease in infants and adults: A systematic review and meta-analysis

**DOI:** 10.64898/2026.02.13.26346212

**Authors:** Alexandra B Hogan, Ainslie Mitchell, Eva Stadler, Yuna Chung, Arnold Reynaldi, Karen M Elias, Steffen Docken, David S Khoury, Timothy Schlub, Miles P Davenport, Deborah Cromer

## Abstract

A number of vaccines and long-acting monoclonal antibodies have been shown to be effective in the prevention of respiratory syncytial virus (RSV) disease. However, an immune correlate of protection for RSV has not yet been identified. We conducted a systematic review to identify published reports of immunogenicity and/or efficacy in vaccines and long-acting monoclonal antibodies against RSV and performed a meta-analysis on extracted data to identify any relationship between antibody increase and protection against RSV disease. We identified 130 relevant reports which we classified into an open access evidence map of RSV immunisation products. We found a strong correlation between the immunisation induced rise in neutralising antibody titres and efficacy (*ρ*>0.7 for all comparisons, Spearman). For infants, we estimated that each 10-fold increase in neutralising antibody titre rise provides an additional 31% [95% CI 10%–47%], 47% [95% CI 36%–56%] and 57% [95% CI 45%–66%] reduction in the relative risk of symptomatic, moderate and severe disease respectively. For older adults, a 10-fold rise in antibody levels was associated with a 34% [95% CI -2%–57%], 50% [95% CI 22%–67%] and 63% [95% CI 36%–79%] reduction in the relative risk of RSV disease with 1, 2 and 3 symptoms respectively. These results align extremely well with findings from natural history studies and individual-based analysis of correlates of protection studies. This work paves the way for use of neutralising antibodies as a correlate of protection to guide the development, approval, and deployment of RSV vaccines and monoclonal antibodies.

## Introduction

Respiratory syncytial virus (RSV) is a common respiratory pathogen. While RSV symptoms are generally mild, with infections occurring throughout the lifespan, it is recognised as a key cause of severe respiratory disease in infants and the elderly^1,2^. Until recently, the only preventative product available for RSV was palivizumab, a high-cost, short-acting monoclonal antibody typically only used in children at heightened risk of serious RSV disease^3^. Over the past 10 years, a number of vaccines and long-acting monoclonal antibodies against RSV have undergone clinical trials, with some now being approved for use as prophylaxis in both infants and older adults^4^. This burgeoning development of RSV-vaccines followed the characterisation of the RSV-fusion (F) glycoprotein in its prefusion conformation in 2013, and identification of a new antigenic site, named Ø^5^.

In 2023, two vaccines were approved: Abrysvo, indicated for people aged 60 years and older and for pregnant women to protect newborn infants in the early months of life; and Arexvy, a vaccine for older adults^6,7^. In addition, in 2022 a new long-acting monoclonal antibody, nirsevimab, was licenced for use (initially in the EU and UK, and subsequently elsewhere) as a single injection in infants following birth, again to provide protection in early infancy^8^. More recently, mRESVIA, a vaccine to prevent RSV use in older adults, was licensed for use in the USA in May 2024, followed by another long-acting monoclonal antibody, Clesrovimab, in June 2025^9,10^.

RSV-specific antibodies have been shown to increase following vaccination or passive antibody administration^11^, however, a thorough examination of both immunogenicity and efficacy conferred by the products en-route to market has not been performed. This means that key questions, such as whether vaccine efficacy is similar for different disease severities and in different populations, remain unanswered. In addition, there is currently no accepted correlate of protection against RSV infection or disease, and while it is assumed that RSV-specific antibodies are important mediators of protection, the relationship between the two has not been quantified. A review of the evidence for correlates of protection against RSV disease in infants found that amongst the studies that investigated correlates of RSV protection, findings were inconsistent^12^.

In this study, we perform a systematic review of the available literature on immunogenicity and efficacy conferred by vaccines and long-acting monoclonal antibodies aimed at protecting otherwise healthy populations against RSV. In addition to analysing the findings from these studies, we combine the results from the identified studies to evaluate the evidence that neutralising antibodies are correlated with protection from RSV disease.

## Results

### Systematic Review and Open Access Evidence Map

We searched PubMed from inception to June 06, 2024 and identified 1460 unique records. After initial screening, we were left with 187 records on which we performed full text review (full details of which can be found in the Supplementary Methods S1.1). We retained 119 reports of studies that investigated RSV vaccines or long-acting monoclonal antibodies and reported RSV antibody and/or efficacy data (Figure S1). An additional 11 relevant reports, identified through reference checking and continued literature surveillance were added, totalling 130 references that report on 76 clinical studies (Figure S1).

We mapped all 130 reports into an evidence map, categorising reports according to the population immunised, the product type administered and the data reported. Given the step change in vaccine development following characterisation of the RSV-prefusion-F protein in 2013, we also categorised reports by whether they were published prior to 2013, or from 2013 onwards, and included this categorisation in our evidence map. This open-access evidence map provides a resource for future studies of RSV vaccine candidates and will be made available following publication.

Of the 130 reports, 53 reported on infants and/or children, 55 reported on general adult or maternal populations, and 43 reported on older adults (≥60 years of age) (Figure 1). Data were presented following both active immunisation (direct vaccine administration, N=112) and passive immunisation (to an infant following vaccination of the mother, N=10, or monoclonal antibody administration, N=16). The majority of documents (112 of 130) reported RSV antibody responses (hereafter referred to as immunogenicity), and 41 reported efficacy or effectiveness against RSV, with 23 reporting both immunogenicity and efficacy (Figure 1). Several different product types were represented in the identified reports, with 16 articles reporting on a long-acting monoclonal antibody, 60 on a protein subunit vaccine, 17 on a viral vector vaccine, 22 on a live-attenuated vaccine, 5 on a combination vaccine, 5 on an mRNA vaccine, 5 on a formalin inactivated vaccine, and 1 on a virus-like particle vaccine. The majority of the reports (105/130) were published in 2013 or later. For the remainder of this work, we focus on these 105 reports published from 2013 onwards (after the characterisation of the pre-fusion F protein), as these represent the current generation of vaccines.

**Figure 1.**
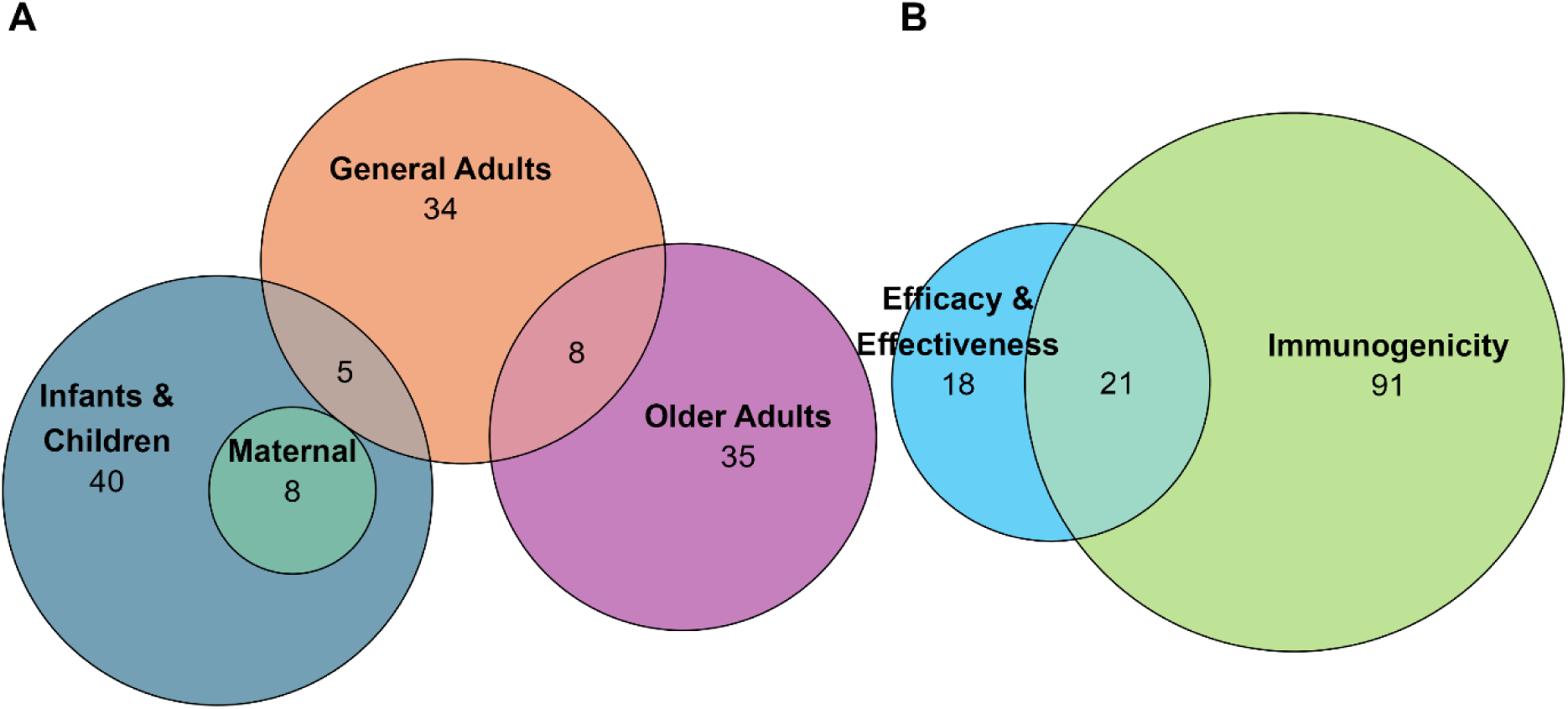
Summary of all reports identified in our systematic review (A) Reports categorised by study population. (B) Reports categorised by data type reported. The age categorisation classifies each report by the population age group in which the product was trialled: either infants (aged ≤1 year); children (aged 1–18 years); general adults (non-pregnant adults aged 18–60 years); maternal (pregnant individuals); and older adults (aged ≥60 years).

### Protection against Natural Infection Afforded by Immunisation

From the 105 relevant documents, we identified 30 reports that provided information from randomised controlled trials (RCTs) on the efficacy of immunising either infants, young children or adults against RSV. Of these, 3 were reports of human challenge studies, and so did not represent protection against naturally acquired infection and therefore were excluded. This left 27 reports of vaccine efficacy (details in Supplementary Table S2), 14 of which were reports of studies conducted in infants or children following either direct immunisation of the infant or child, or immunisation of the mother prior to birth, and 13 were conducted in older adults. Of the 27 reports, 14 either reported repeat data or aggregated efficacy across different dose regimens, leaving 13 reports that reported unique efficacy information for a defined formulation (7 in infants and children, and 6 in older adults). These data are summarised in Figure 2A and B.

**Figure 2.**
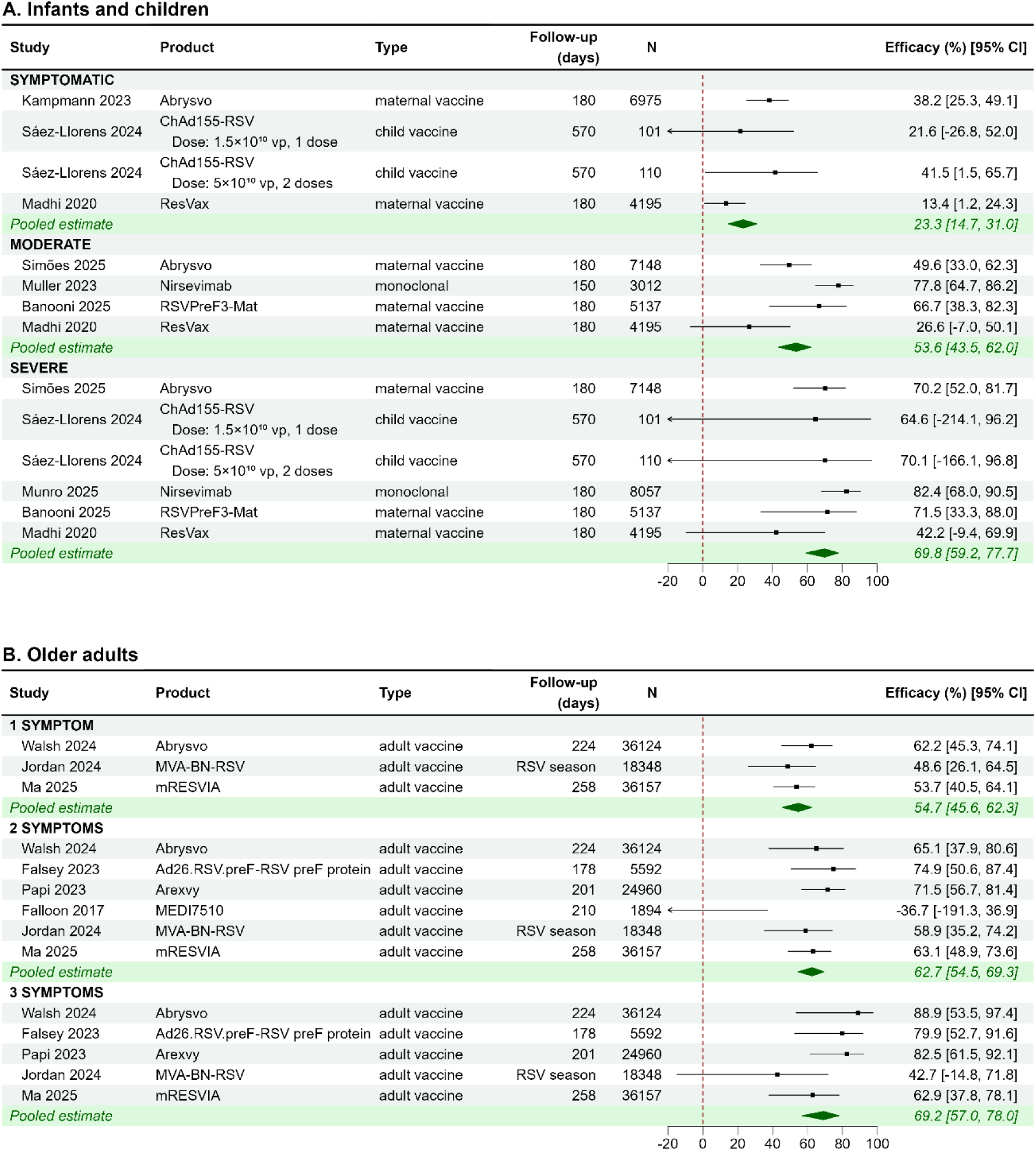
Reported efficacy conferred from immunisation of (A) infants and children (7 reports) and (B) older adults (6 reports) for products identified in the systematic review. Data are shown for each severity level reported in the primary reports, and efficacy is calculated as 1 – RR, where RR is the unadjusted relative risk calculated from the raw data in each report. Where protection over several follow-up periods was captured within one report, we show the protection estimate documented for the period closest to 6 months (180 days). N represents the total at-risk population in the study (across placebo and intervention groups) to indicate the study size. vp: viral particles.

Protection was reported against different endpoints in infants and older adults. Adult endpoints were defined based only on the number of reported symptoms, however infant and child endpoints were determined by both the symptoms themselves and interventions required (Supplementary Methods S1.4). We classified outcomes into three distinct severity categories. For infants, severity was classified as either symptomatic, moderate, or severe, and for adults, severity was categorised into outcomes with either 1 symptom, 2 symptoms, or 3 symptoms.

Across all products and populations, there was clear evidence that vaccination afforded protection against RSV disease of all levels of severity, with 12 of the 13 references (92%) reporting a positive efficacy against all severity outcomes (Figure 2).

We found a consistently higher level of protection against more severe outcomes in both infants and adults, though this difference was more evident in the outcomes presented for infants (Figure 2A). In infants, the pooled efficacy estimate was 23.3% [95% CI 14.7–31.0] against symptomatic outcomes versus 53.6% [95% CI 43.5–62.0] and 69.8% [95% CI 59.2–77.7] against moderate and severe outcomes respectively. In older adults the pooled efficacy estimates were 54.7% [95% CI 45.6–62.3], 62.7% [95% CI 54.5–69.3] and 69.2% [95% CI 57.0–78.0] against RSV disease with 1, 2 or 3 symptoms, respectively. Pooled efficacy estimates are also given in Supplementary Table S9.

### Impact of Immunisation on Antibody Titres

We next sought to ascertain which reports within our systematic review reported on antibody levels following immunisation with any of the products shown in Figure 2. We identified 35 documents that reported unique data on antibody titres following immunisation with the same formulation (matching dose and adjuvant) that was reported in an efficacy or effectiveness study. These are detailed in Supplementary Table S3. Nine reports included data from infants or children, 8 from general adults, 6 from maternal populations and 18 from older adults. We used these data to calculate the maximum difference in neutralising antibody concentration and IgG titre between the immunised and placebo cohorts, following either administration (for immunised individuals) or birth (infant blood levels, in cases where the mother was immunised). This difference is hereafter referred to as the fold-rise. For details on how the fold-rise in antibody titre was calculated see Supplementary Methods S1.5. Neutralising antibody rises are shown in Figure 3, for infants and children (panel A), general adults (B), maternal populations (C), and older adults (D). Supplementary Figure S3 shows the corresponding fold-rises in IgG titres.

**Figure 3.**
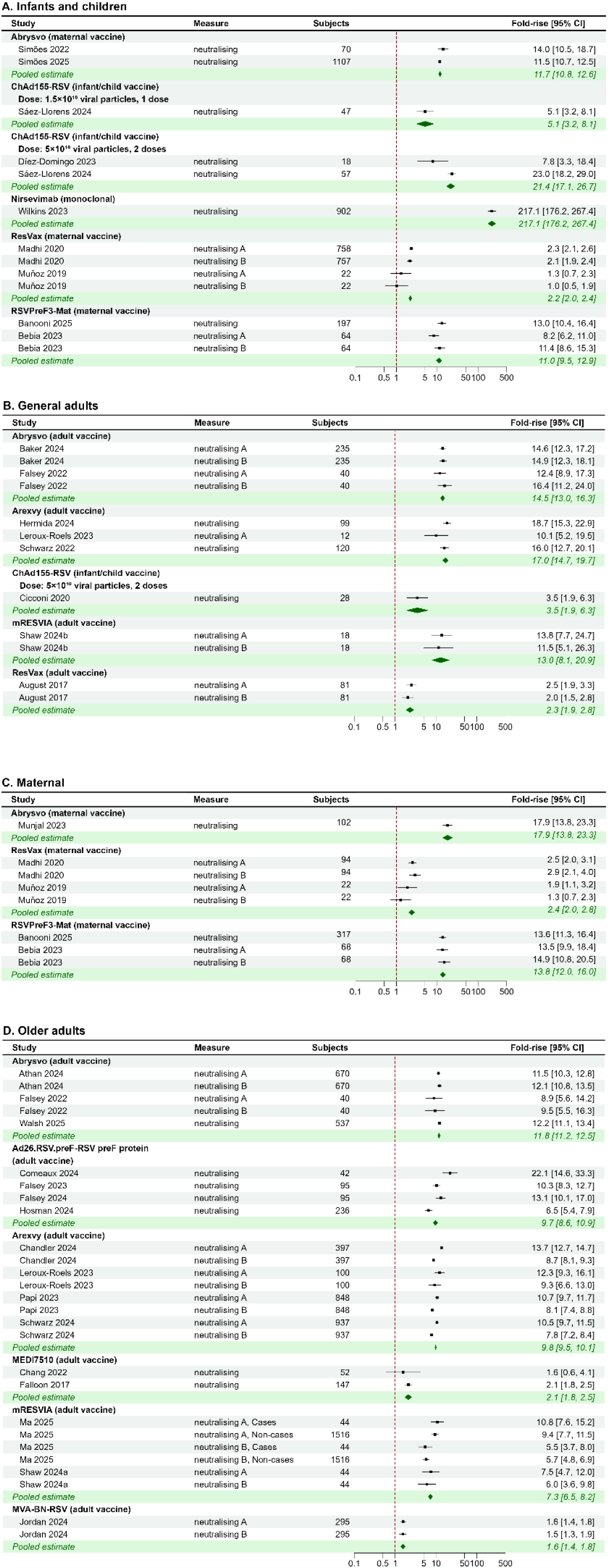
Neutralising antibody fold-rise for different RSV immunisation products in (A) infants and children (9 reports); (B) general adults (8 reports); (C) maternal populations (5 reports); and (D) older adults (16 reports). Individual published reports are grouped by immunisation product, with the pooled estimate for each product shown on the green shaded rows.

As expected, since these products were all taken forward to efficacy trials, all showed rises in both neutralising and binding antibody titres. However, the magnitude of the estimated rise varied significantly from under 2-fold to over 200-fold. This difference may be due in part to the different assays that were used in the different studies but may also reflect the underlying immunogenicity of the products involved. Of the 35 reports in which we assessed antibody titres, 23 reported information for both neutralising and binding antibody titres. We found that out of the 29 study populations in which a comparison was possible, 24 (83%) showed a greater rise in binding antibody titres as compared to neutralising antibody titres (p=<0.001, two-sided Wilcoxon signed-rank test). This may reflect the different assays used to measure neutralising and binding antibody titres.

### Does the Rise in Antibody Titre Correlate with Efficacy?

Given our findings that almost all products showed evidence of protection and all showed a rise in antibody titres, we asked whether the magnitude of the rise in antibody titres following immunisation was correlated with the efficacy afforded. To do this, we linked each of the efficacy values in Figure 2 with the corresponding antibody fold-rises from Figure 3, for the same product, formulation, and population group. We were able to do this for five products in infants and children: Nirsevimab (AstraZeneca & Sanofi)^13,14^, RSVPreF3-Mat (GSK)^15^, ResVax (Novavax)^16^, ChAd155-RSV (GSK)^17^, and Abrysvo (Pfizer)^7,18^ and six products in adults: MVA-BN-RSV (Bavarian Nordic)^19^, Arexvy (GSK)^20^, Ad26.RSV.preF–RSV preF protein vaccine (Janssen)^21^, MEDI7510 (MedImmune)^22^, mRESVIA (Moderna)^23^, and Abrysvo (Pfizer)^24^. Where there were multiple immunogenicity reports corresponding to a single efficacy measurement, we calculated a weighted average of all reported values (methods described in Supplementary Methods S1.5). Included reports are detailed in Supplementary Tables S4 and S5.

We observed a strong positive association between the fold-rise in antibody levels and clinical protection across the three reported outcomes in children (Spearman *ρ*=1 for symptomatic and moderate disease and *ρ*=0.8 for severe disease), noting that these findings should be interpreted cautiously due to the small number of points (Figure 4A). There was a similar strong association between the fold-rise in antibody levels and clinical protection in adults (Spearman *ρ*=1 for 1 and 3 symptoms, *ρ*=0.71 for 2 symptoms), with a significant correlation for the outcome of 3 symptoms, despite the small number of points (Figure 4B).

**Figure 4:**
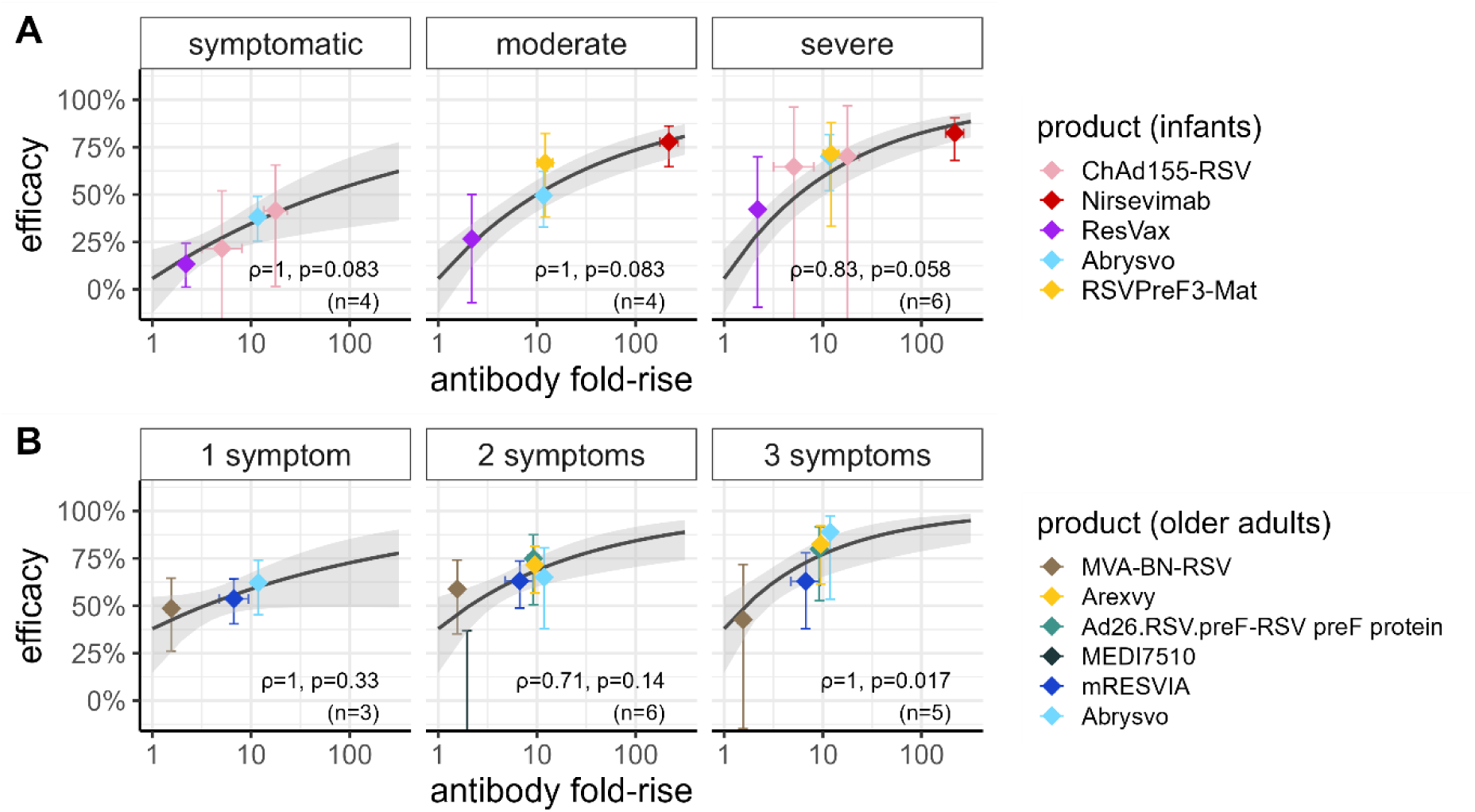
Modelled protection against RSV in (A) infants and children and (B) older adults. Relationship between the fold-rise in neutralising antibody titre and efficacy in (A) infants, against symptomatic, moderate, and severe RSV; and in (B) older adults, against RSV with 1, 2, or 3 symptoms. The immunisation products are shown as coloured diamonds with corresponding confidence intervals. In each panel, the black line represents the fitted statistical model, and the grey shaded region represents the 95% confidence interval. For each of infants and children, and older adults, the fitted model is a model with a single fitted intercept, and a fitted slope for each severity level (Table S6, “Varying slope model”). On each panel, the annotated text reports the Spearman’s rho and p-value, measuring the non-parametric correlation between the antibody fold-rise and efficacy for each population group and severity outcome combination. We note that due to the small number of points in each comparison, we would not expect significance in the p-values, however they are reported for completeness. The models fitted to the data expressed as the relative risk are shown in Figure S4.

In order to better quantify the relationship between rises in antibody titres and efficacy in these populations, we fit two linear mixed effects models to the datasets of eligible matched immunisation formulations, one for infants and one for older adults. The models related the rise in antibody titres following immunisation to the relative risk of RSV disease in infants and in older adults (Supplementary Table S6), and included two key parameters, namely a slope and an intercept. They also included a random effect parameter to capture the report from which the efficacy data was derived. Full details of the models are described in Supplementary Methods S1.6. The slope identifies the extent to which neutralising antibodies are correlated with protection and the intercept indicates the amount of protection that could potentially be conferred by non-antibody-mediated effects. We fit the models to all severity outcomes simultaneously but generated separate models for infants and adults.

We initially allowed the potential relationship between antibodies and protection to be the same for each severity outcome (but different for infants and adults). We found evidence that neutralising antibodies were correlated with protection in both infants and adults (F-test comparing a model with no slope parameter gives *p<*0.001 and *p=*0.018 for infants and adults respectively). Additionally, there was no evidence of an increase in protection without an associated increase in antibody levels for either infants or adults (F-test comparing to a model with no intercept parameter gives *p*=0.38 and *p=*0.29 for infants and adults, respectively).

Next, given the observed higher efficacy following immunisation against more severe outcomes in both infant and older adult populations (Figure 2), we tested whether allowing the relationship between antibodies and protection to vary for different severity outcomes improved the model fit in either population (i.e., we allowed the slope parameter to vary by severity levels, Figure 4A). In infants, the model that included three different slopes provided a significantly better fit to the data (*p*=0.004, F-test). We estimated that each 10-fold increase in neutralising antibody titre rise reduces the relative risk of symptomatic disease by 31% [95% CI 10%–47%] and decreases the relative risk of moderate and severe disease by 47% [95% CI 36%–56%] and 57% [95% CI 45%–66%], respectively.

In older adults, there was a similar trend towards an increase in antibody titre providing greater protection against more severe outcomes, though this was less significant than in infants. A model that allowed for different slopes for outcomes with different numbers of symptoms provided a slightly better fit to the adult data (*p*=0.045, F-test). We estimated that each 10-fold increase in neutralising antibody titres reduces the relative risk of RSV disease with 1, 2 and 3 symptoms by 34% [95% CI -2%–57%], 50% [95% CI 22%–67%] and 63% [95% CI 36%–79%], respectively, in adults (Supplementary Table S9).

In order to confirm that the findings above were not dependent on the choice of model structure or data choice, we performed two separate sensitivity analyses. These were: (i) Using an alternative method to calculate the fold-rise in neutralising antibody titre (Figure S2), and (ii) Incorporating uncertainty in the estimate of the fold-rise in antibody titres into our model fitting (i.e., x-axis uncertainty, Supplementary Methods S2.2). Each of these sensitivity analyses confirmed our finding that neutralising antibody titre is correlated with protection against RSV disease in both children and adults.

### Relationship to Existing Therapeutics and Natural History Studies

Having established evidence for a relationship between the rise in neutralising antibodies and protection from RSV disease, we sought to determine whether this relationship aligned with that observed in studies using existing therapeutics in other populations, in natural history studies, and in analysis of individual correlates of protection studies.

#### Existing Therapeutics

Monoclonal antibodies with relatively short half-lives have long been available for use in populations not included in our systematic review – namely, in preterm infants, and those at high risk of RSV-related complications. These antibodies are administered intravenously multiple times over an RSV season, and have been estimated to provide a 7-fold [95% CI 5–9] to 12-fold [95% CI 6–21] rise in neutralising antibody titres at one month after the first dose^25,26^. A recent Cochrane review estimated the effectiveness of palivizumab against hospitalisation as 56% [95%CI 36–70]^3^. As can be seen in the right most panel of Figure 5A, this aligns almost exactly with our estimate of the protection expected in infants for such a rise in neutralising antibody titres.

**Figure 5:**
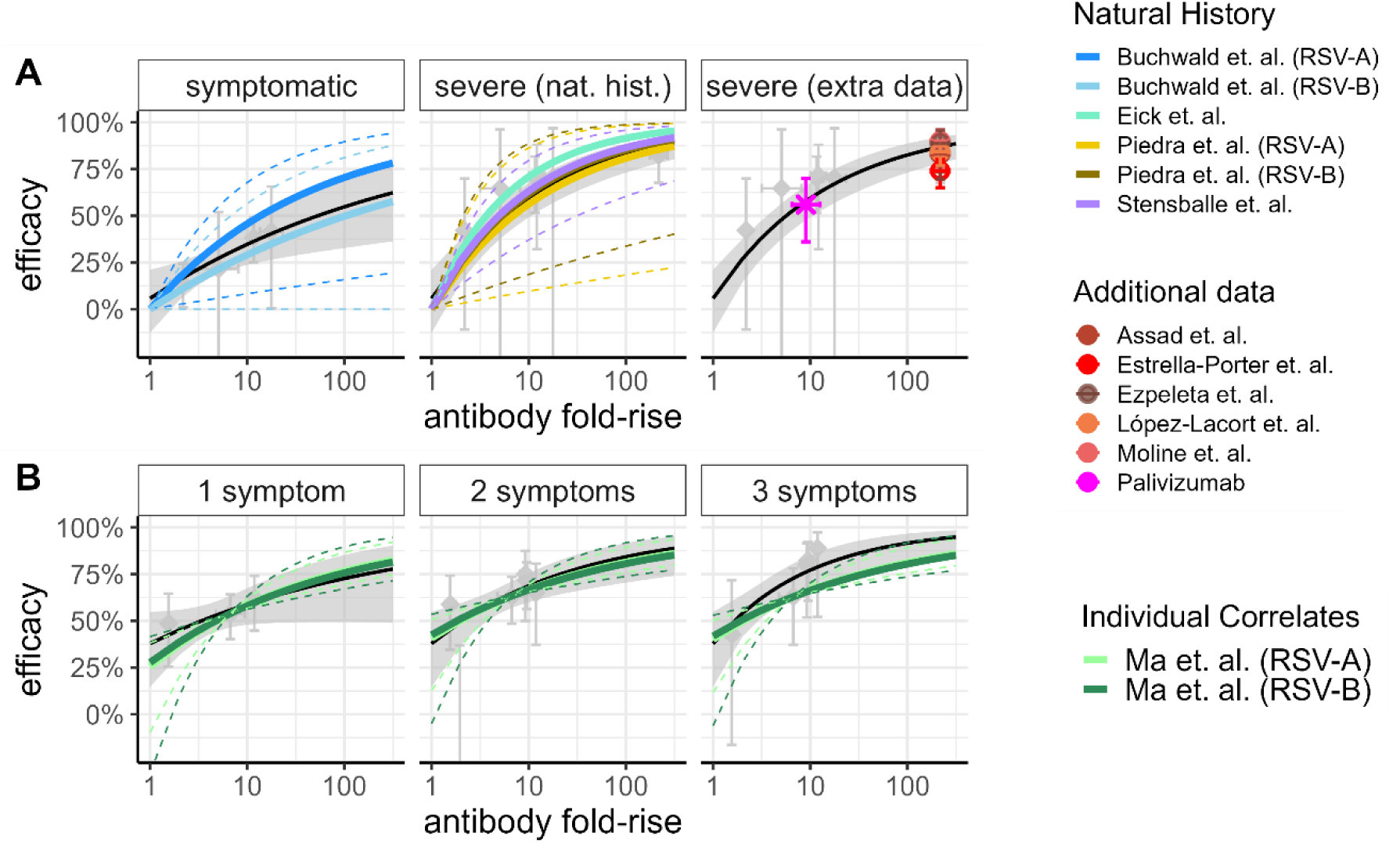
Agreement between our model and estimates of the relationship between neutralising antibodies and protection found in the literature. Agreements are shown for both infants and children (panel A) and older adults (panel B). In each panel, the solid black line represents the fitted statistical model (the “varying slope model”) depicted in Figure 4, and the shaded black ribbons show the 95% confidence intervals of the fit. Coloured points and lines show estimates from the literature (A) Infants and children: The coloured solid lines in the left two panels illustrate estimates from four studies of the relationship between infection-induced neutralising antibody titres and protection from RSV disease and/or hospitalisation in infants^32–35^. Dashed lines show the corresponding 95% confidence intervals. The coloured circles in the right most panel show estimates of antibody rises and protection for nirsevimab found in the literature. The magenta circle and confidence intervals show the pooled estimate of the efficacy of the monoclonal antibody, palivizumab, against hospitalisation matched to the estimated geometric mean of the fold-rise in neutralising antibody titres following palivizumab administration^3,25,26^. The red circle and confidence intervals show the estimate of nirsevimab efficacy from a case control study matched against the estimated fold-rise in antibody titres following nirsevimab administration^25,27^. (B) Older adults: The light and dark green solid lines illustrate the estimated relationship between antibody fold-rise with protection against RSV disease for the mRESVIA vaccine product^23^. Dashed lines show the corresponding 95% confidence intervals. Note that these lines are anchored at the estimated protective efficacy of the mRESVIA vaccine product against RSV disease with 1, 2 or 3 symptoms, respectively.

#### Observational Studies

Within our systematic review, we identified six reports of observational studies of nirsevimab protection in infants, five of which reported an adjusted effectiveness measure. We sought to determine whether the effectiveness reported in those studies aligned with the efficacy predicted by our model for the rise in antibody titres conferred by nirsevimab administration. The identified reports of observational studies reported adjusted effectiveness estimates of 74%–90% against hospitalisation^27–31^, which align very closely with the efficacy of 86.8% [95% CI 78.1–92.1] predicted by our model (Figure 5A, right panel).

#### Natural History Studies and Individual Correlates of Protection Studies

A number of studies have been conducted that correlate neutralising antibodies with protection from RSV disease within a cohort. For infants, we identified one study that estimated the association between (naturally occurring) antibody titres and protection from any RSV disease^32^, and three that reported on the relationship between RSV antibodies and protection from hospitalisation for RSV^33–35^. For adults, we identified one paper, reporting on Moderna’s mRNA-1345 (mRESVIA) RSV vaccine product, that correlated an individual’s rise in neutralising antibody titres with the increase in protection^23^.

We identified only one study in infants that related antibody titres to protection from RSV infection, and this study reported that a 10-fold increase in antibodies was associated with a 29–46% reduction in the odds of RSV infection (depending on whether the antibodies were targeting RSV-A or RSV-B subtypes)^32^. This is exactly in line with estimates from our model, which predicts a 31% [95% CI 10%–47%] reduction in the risk of symptomatic RSV disease for a similar antibody rise (Figure 5A, left panel). Two other identified studies considered cord blood titres in infants and estimated a 63% and 71% decrease in the risk of hospitalisation for RSV for each 10-fold increase in RSV-associated neutralising antibody levels^33,35^, while a third study that measured antibody titres on hospital admission, reported a 56–62% decrease in the odds of hospitalisation for each 10-fold increase in antibody titres^34^ (depending on whether the antibodies were targeting RSV-A or RSV-B subtypes). These estimates are once again in line with our estimate of a 57% [95% CI 45%–66%] decrease in the risk of severe disease for each 10-fold increase in neutralising antibody levels (Figure 5A, middle panel).

In the correlates of protection study of adults, a 10-fold increase in vaccine-induced neutralising antibody titre was reported to be associated with a 45–59% decrease in the hazard ratio of disease (depending on RSV strain and outcome severity)^23^. This once again aligns with the 34%–63% decrease in the risk of disease estimated by our model (Figure 5B). Interestingly, the study by Ma et. al. in adults did not report a greater reduction in the hazard ratio against more severe outcomes as compared to milder outcomes for the same increase in neutralising antibody titre^23^.

Thus, we can conclude that our estimate of the relationship between neutralising antibody rises and protection from disease is consistent with real-world evidence.

### Predicting Vaccine Efficacy in Younger Populations

RSV vaccines designed to protect adults are currently approved for use in populations over 60 years of age in the US, UK and Australia (amongst other countries). An important question is how these vaccines will perform in younger populations in whom vaccine efficacy has not yet been evaluated. The relationship between vaccine immunogenicity and efficacy demonstrated in this study allows us to predict vaccine effectiveness in different populations based on observed immunogenicity. We find that vaccination of adult cohorts under 50 years of age results in rises in neutralising antibody titres that are between 1.2 and 1.8-fold greater than those observed in adults over 60 years of age (Abrysvo: 1.2-fold [95% CI 1.1–1.4], Arexvy: 1.7-fold [95% CI 1.5–2.0], mRESVIA: 1.8-fold [95% CI 1.1–2.9], Supplementary Table S10).

Using the correlation between neutralising antibody rises and efficacy presented in Figure 4B for older adults, we can estimate the impact of this greater rise in neutralising antibody titres on the relative risk of disease (assuming the same relationship also holds for younger adults). We predict that a 20–80% increase in the rise in antibody titres following vaccination in younger adults will result in an additional 1.03–1.29-fold reduction in the relative risk of disease following vaccination. Our modelling suggests that the greatest reduction in relative risk is observed against more severe disease. This translates into a predicted 1–7% increase in the observed vaccine efficacy over the efficacy ranges observed in practice. Thus, combining both the meta-analysis of neutralising antibody titres and the immunogenicity data in younger adults suggests that vaccines are likely to afford similar, or slightly higher efficacy in younger populations than that observed for older populations.

## Discussion

In recent years, a number of vaccines and long-acting monoclonal antibodies have been licensed for use for RSV prophylaxis in both adults and infants. Although the effectiveness of passive antibody therapy indicates an important role for antibodies in protection, a correlate of protection relating immunogenicity with efficacy against RSV disease has not yet been established. One challenge in studying antibody levels and protection in RSV is the background level of antibodies in the population due to natural exposure (which varies with age). For this reason, we have chosen to quantify the immune effect of an intervention as the ‘fold-rise’ in antibody levels in the treated versus control populations. Our study includes data from RCTs of 10 products and integrates this with studies comparing the rise in antibody titres in immunised individuals compared to controls. We find a strong association between the fold-boost in antibody levels and protection across different products, populations (adult vs. infant), and clinical outcomes.

Across all vaccine and monoclonal antibody studies, the reported efficacy was higher against severe outcomes than against mild or moderate outcomes, and we found evidence that a similar increase in antibody titres provides a greater increase in protection against more severe outcomes. This was true regardless of the population (infants or older adults) or the immunisation product being considered. Thus, for prophylactic products, improving the boost to antibody titres is likely to have a greater impact on protection against severe disease than against symptomatic disease. The vast majority of the products in both infants and adults conferred rises in antibody titres of around 10-fold, and those identified as having been taken through to market all reported a fold-rise of at least 7-fold (Figure 3). The monoclonal antibody, nirsevimab, reported the highest rise in antibody titres of around 100-fold.

One of the strengths of our study is the inclusion of all available data from vaccine and long-acting monoclonal antibody products, formulated to protect both infants and adults against RSV. By doing this we have provided a comprehensive overview of the prophylactic landscape for RSV and generated an open access evidence map that can be used by researchers in the field. Combined, these data suggest that the fold-boost in antibody levels from an intervention is a robust predictor of the efficacy of that intervention. However, the inclusion of diverse studies also provides a number of limitations. Firstly, there were differences in baseline levels of immunity, products and studies between the different populations we considered. In infants, both pre-existing immunity and immunity conferred from four of the five products analysed was passively acquired, while in adults immunity was actively acquired (from prior infection or direct vaccination). Thus, in addition to the differences in age, there is also a difference in the type of immunity being considered between these two populations.

Secondly, antibody titres analysed in this analysis were all measured using different in vitro assays, with different sensitivities and dynamic ranges. Although we have attempted to account for these assay differences by considering only the fold changes in antibody titres, rather than the raw titres themselves, we cannot discount the possibility that variations in assay design across the different studies could affect the estimates of the fold-rise in antibodies. However, we note that for each individual product within a given population, the estimated fold-rise in antibodies was calculated by incorporating data from a median of 2 separate in vitro studies (range 1–4), likely reducing the effects of assay variability. In addition, we compared our predicted relationship between antibodies and efficacy with the relationship derived from four studies of naturally occurring antibodies and protection in infants. These natural history studies each used a single assay (within each study) to measure neutralising antibody levels across the population, and correlate this with risk. The similarity between these associations and our curve derived from the data from multiple different studies using a range of in vitro assays give confidence that our estimate was not severely affected by assay variability. Finally, a high variability in estimates between assays would have been more likely to mask a potential correlation between antibodies and protection rather than enhance it, and therefore we believe that assay differences should not impact the conclusions drawn in this work.

Finally, the reported clinical outcomes across studies are quite different. For infants, efficacy is reported against outcomes that primarily rely on symptom severity, and often relate to pre-specified thresholds for oxygen saturation and respiratory rate. Although these may differ for different studies, by and large there is broad agreement between disease outcomes that can be classified as “symptomatic”, “moderate” and “severe” disease in infants (Supplementary Methods S1.4). In contrast, adult efficacy studies report outcomes against disease with different numbers of symptoms (usually one, two or three symptoms). This means that reported “efficacies” in the infant and adult populations often have different meanings, and it is difficult to address the question of whether a certain product (or certain rise in antibody titres) has a better protective efficacy in infants or adults. Were such a comparison to be of interest, guidelines would need to be developed that define comparable disease severities in infants and adults.

Another limitation of our work is that the reported protection against multiple different outcomes for a given product was often taken from the same study, meaning that these estimates were not independent of each other. Availability of matched immunogenicity and efficacy data (i.e. obtained from the same cohort) or from additional products would increase certainty in the model results.

The correlation between antibodies and protection described here clarifies some of the questions raised by an earlier review of the correlates of protection for RSV in infants^12^, and aligns with a study analysing individual correlates of protection in adults^23^. In addition, our estimate of the protection gained by a rise in neutralising antibody titres aligns almost exactly with the estimates available in natural history studies, and in studies of existing, short-acting monoclonal antibodies offered to pre-term and at-risk infants (Figure 5).

Importantly, our work does not provide a single ‘threshold’ value for the antibody level that provides clinical protection. Instead, we show that protection varies with the fold-rise in antibodies in the treated population (which is dependent on the baseline antibody level). In addition, the fold-rise associated with a given level of protection varies slightly by population and clinical severity (Figure 4). This is consistent with results in COVID-19, where there is an established relationship between neutralising antibody titre and protection which varies by clinical severity^36,37^. In addition, we note that although for influenza a hemagglutinin inhibition assay (HAI) titre of 40 is often regarded as a ‘threshold of protection’, the relationship between HAI titre and protection in fact shows a gradual increase in protection with titre^38,39^, similar to the relationship between antibody rise and protection described here for RSV.

The strength of the association we have uncovered between the fold-rise in antibody titres and protection against RSV across populations and outcomes suggests that rises in neutralising antibody titres are an effective means by which to demonstrate non-inferiority with existing products, thereby facilitating product approval and licensure processes. It also provides a way to extrapolate vaccine effectiveness predictions to populations beyond those originally studied (e.g. in younger adult populations or immunocompromised individuals) by evaluating the rise in neutralising antibody titres and using this to predict vaccine efficacy. In addition, it supports the use of this correlate to provide scientific insights around planning of product delivery and prioritisation (for example, by helping estimate protection and thereby informing epidemiological and transmission models as well as cost-effectiveness analysis under different scenarios). Thus, the correlate of protection derived in this work will aid in development, testing, assessment, and deployment of future interventions designed to provide protection against RSV.

## Methods

### Search Strategy and Data Extraction for Efficacy and Immunogenicity Studies

We searched PubMed for publications of primary studies reporting on RSV immunogenicity and efficacy against RSV disease in healthy individuals, following immunisation with an RSV vaccine or monoclonal antibody, or following birth in infants born to RSV-immunised mothers, published from database inception to June 06, 2024. We identified additional reports through searches of grey literature and by reviewing references cited in news and other non-primary research articles that were eligible for full-text review. Publications presenting updated trial results for the identified products were included up to August 2025. Full details of the search strategy and implementation are given in the Supplementary Methods S1.1.

For each report, we extracted information on the product, population, and the number of subjects. For reports of effectiveness studies, we additionally extracted the number of subjects in the immunised and placebo groups, the number of events in each group, the outcome measures, and the study period. For reports of immunogenicity studies, we also extracted reported antibody titres, either RSV neutralising titres or anti-RSV IgG. Full details of the data extraction methods are provided in the Supplementary Methods S1.2.

### Categorisation of Studies

We categorised the individuals in each study by age class, defined as follows: infants (younger than one year); children (aged one to 18 years); general adults (18 to 60 years); maternal (pregnant individuals); and older adults (older than 60 years). The efficacy studies captured two age groups, infants and older adults, and we assigned different levels of severity based on the medical outcome classification definitions used in these studies (see Supplementary Methods S1.4). For infants, severity was classified as either symptomatic, moderate, or severe, and for older adults, severity was categorised by symptoms, into outcomes with either 1 symptom, 2 symptoms, or 3 symptoms.

### Selecting Efficacy Studies for Analysis

Studies identified in our review reported efficacy at multiple timepoints that varied by study; these could be from 90 days through to longer than two years. For the purposes of correlating antibodies with protection, we were interested in protection over the first RSV season following vaccination, with enough follow-up time to allow for a reasonable estimate of vaccine efficacy. Therefore, we only incorporated efficacy at a single time point from each report, and where efficacy was available at multiple time points, we included the estimate that was estimated closest to six months (180 days). Where multiple documents reported efficacy estimates for the same cohort (for example due to interim and final analyses), we only incorporated data from the most recent report. Where, in addition to reporting efficacy against RSV, a document reported efficacy against RSV-A and RSV-B strains separately, we used the combined measure (against both strains) only. Finally, if a report did not report efficacy against an outcome that fit into one of our severity categories, it was not included in further analysis (this applied to only one report)^41^.

### Selecting Immunogenicity Studies for Analysis

Having identified the efficacy studies that could be used in our analysis, we next identified the immunogenicity studies that reported antibody titres following administration of a corresponding product to the same age group, at the same dose, and with the same adjuvant. This enabled us to match each efficacy outcome with a corresponding immunogenicity measure.

### Estimating the Rise in Antibody Titre Following Immunisation

We calculated the rise in antibody titre following immunisation as the difference between the mean neutralising antibody titres (log_10_ scale) in immunised and placebo groups. This difference was calculated either at birth (for infants whose mothers had received a maternal vaccine) or at the first measurement taken more than a week after immunisation (for subjects receiving monoclonal antibodies or a vaccine). The 95% confidence interval on the rise in antibody titres was calculated using the reported confidence intervals on each antibody concentration measure in the reports and the formulae are given in the Supplementary Methods S1.5. Where titres against RSV-A and RSV-B were reported separately, we calculated the overall rise in neutralising antibody titre as the geometric mean of the titres against the two strains (see Supplementary Methods S1.5).

### Modelling the Relationship Between Antibodies and Efficacy

We fitted a linear mixed effects meta-regression model to estimate the relationship between RSV-specific antibodies and the relative risk of RSV disease for each of infants and older adults, with fixed effect terms for the slope and the intercept, and a random effect to account for variability between studies. We also tested models with additional terms to allow for the correlation to differ by severity category, by allowing either the slope or intercept to vary by severity category. The model was fitted using the rma.mv() function with maximum likelihood estimation, from the metafor package version 4.8-0 in R version 4.5.0^42^. Full details of the model are given in the Supplementary Methods S1.7.

We additionally tested the sensitivity of the fitted model parameters to an alternative calculation of the fold-rise in neutralising antibody titre in infants, and finally used multiple imputation combined with Rubin’s Rules to estimate the sensitivity of results to uncertainty in the fold-rise in antibody titre (Supplementary Methods S2).

## Data Availability

All data, code, and the evidence map will be made available on GitHub upon publication.

## Acknowledgements

We would like to acknowledge helpful discussions with Shanchita Khan and Bronwyn Bailey.

## Funding

ABH, ES, DSK, MPD and DC are funded by Australian National Health and Medical Research Council Investigator Grants (Numbers 2009278 (ABH), 2034282 (ES), 2033318 (DSK), 2034108 (MPD) and 2026360(DC)). DSK and MPD are funded by the Medical Research Future Fund (MRFF, Australia) (MRFF 2016062 (DSK, MPD) and MRFF 2015313 (MPD)). KME is funded by an Australian Government Research Training Program (RTP) Scholarship. The contents of the published material are solely the responsibility of the authors and do not reflect the views of the funding bodies.

## Supplementary Material

### 1. Supplementary Methods

#### 1.1. Systematic Review

A systematic search of the literature was performed using PubMed from database inception to June 06, 2024.

Citations identified from the literature searches were imported to Covidence (covidence.org). Titles and abstracts of articles were manually screened in Covidence by two reviewers independently (AM and YC), and conflicts were discussed until consensus was reached. If necessary, a third reviewer (DC or ABH) was consulted for a final decision. Duplicates were removed manually. Full texts of all potentially eligible articles (and those for which eligibility was uncertain) were retrieved. Next, two reviewers (AM and YC) independently reviewed full-text articles in Covidence for inclusion, using the inclusion and exclusion criteria outlined below. Again, conflicts were resolved by discussion and consensus, with a third researcher (DC or ABH) consulted to make a final decision if needed. Citations that failed to meet the inclusion criteria were excluded, and the exclusion reasons were recorded. Additional reports were identified by manual searching of reference lists of identified articles and an ongoing review of relevant literature.

Inclusion and exclusion criteria were based on the PICOS framework (Population, Intervention, Comparator, Outcomes, and Study type).

- Population: We included reports capturing individuals of any age or gender. We excluded studies that specifically focused on premature and immunocompromised populations, high-risk populations with pulmonary conditions, and RSV-infected populations.
- Intervention: We considered all RSV vaccines and long-acting monoclonal antibodies. Studies of RSV short-acting monoclonal antibodies (e.g., palivizumab) or immunoglobulin therapy were excluded because these products are primarily given to premature infants and/or immunocompromised populations.
- Comparator: Studies needed to include either a placebo, control, or standard care group, or have a baseline measure recorded in the intervention group.
- Outcomes: To be included, documents had to report either (i) RSV neutralising titres or anti-RSV IgG/IgA antibody levels in serum, plasma, or nasal fluids for at least one time point following birth or administration of the RSV vaccine or monoclonal antibody; or (ii) vaccine or monoclonal antibody efficacy or effectiveness against RSV infection or RSV disease evaluated over any follow-up period. Reports that solely presented anti-G or N protein antibody data, anti-RSV IgG/IgA subclass data, or anti-RSV IgM data, or that presented exclusively efficacy data from phase 1 trials (or with fewer than 25 participants in the vaccine or placebo study arms) were excluded.
- Study type: We included clinical trials and observational studies. Preclinical studies and non-primary studies, e.g., reviews, were excluded.
- Report type: Published and unpublished reports were included. There was no restriction on publication date. Non-English articles were excluded during screening.

No systematic review protocol was pre-registered for this review.

The search string for this systematic review is listed below. Reports of animal studies were removed by using the Cochrane humans filter and an approach developed by the authors that used mouse, monkey, and bovine-related terms. To exclude reviews, we applied the PubMed ‘review’ and ‘systematic review’ filters to the search strategy. No language limits were applied.

Search String:

((“Immunogenicity”[Title/Abstract] OR “antibod*”[Title/Abstract] OR “efficac*”[Title/Abstract] OR “effectiveness”[Title/Abstract] OR “protect*”[Title/Abstract] OR “Antibodies”[MeSH Terms:noexp]) AND (“respiratory syncytial virus”[Title/Abstract] OR “Respiratory Syncytial Viruses”[Title/Abstract] OR “RSV”[Title/Abstract] OR “Respiratory Syncytial Virus Infections”[MeSH Terms] OR “respiratory syncytial virus, human”[MeSH Ters]) AND (“vaccin*”[Title/Abstract] OR “monoclonal*”[Title/Abstract] OR “Vaccines”[MeSH Terms:noexp] OR “Vaccine Efficacy”[MeSH Terms] OR “immunogenicity, vaccine”[MeSH Terms] OR “Respiratory Syncytial Virus Vaccines”[MeSH Terms] OR “antibodies, monoclonal”[MeSH Terms:noexp])) NOT ((“animals”[MeSH Terms] NOT “humans”[MeSH Terms]) OR (“calf”[Title/Abstract] OR “cow”[Title/Abstract] OR “cows”[Title/Abstract] OR “cattle”[Title/Abstract] OR “bovine”[Title/Abstract] OR “BRSV”[Title/Abstract] OR “calves”[Title/Abstract] OR “mouse”[Title/Abstract] OR “mice”[Title/Abstract] OR “murine”[Title/Abstract] OR “monkey”[Title/Abstract] OR “monkeys”[Title/Abstract]) OR ((((“Immunogenicity”[Title/Abstract] OR “antibod*”[Title/Abstract] OR “efficac*”[Title/Abstract] OR “effectiveness”[Title/Abstract] OR “protect*”[Title/Abstract] OR “Antibodies”[MeSH Terms:noexp]) AND (“respiratory syncytial virus”[Title/Abstract] OR “Respiratory Syncytial Viruses”[Title/Abstract] OR “RSV”[Title/Abstract] OR “Respiratory Syncytial Virus Infections”[MeSH Terms] OR “respiratory syncytial virus, human”[MeSH Terms]) AND (“vaccin*”[Title/Abstract] OR “monoclonal*”[Title/Abstract] OR “Vaccines”[MeSH Terms:noexp] OR “Vaccine Efficacy”[MeSH Terms] OR “immunogenicity, vaccine”[MeSH Terms] OR “Respiratory Syncytial Virus Vaccines”[MeSH Terms] OR “antibodies, monoclonal”[MeSH Terms:noexp])) NOT ((“animals”[MeSH Terms] NOT “humans”[MeSH Terms]) OR (“calf”[Title/Abstract] OR “cow”[Title/Abstract] OR “cows”[Title/Abstract] OR “cattle”[Title/Abstract] OR “bovine”[Title/Abstract] OR “BRSV”[Title/Abstract] OR “calves”[Title/Abstract] OR “mouse”[Title/Abstract] OR “mice”[Title/Abstract] OR “murine”[Title/Abstract] OR “monkey”[Title/Abstract] OR “monkeys”[Title/Abstract]))) AND (“review”[Publication Type] OR “systematic review”[Filter])))

#### 1.2. Data Extraction and Collation

Data were extracted from each eligible report into a bespoke form by one reviewer (AM or YC). Extracted data were subject to spot checks by a second reviewer (AM or YC, approximately 65% of the data), and any disagreements were discussed until a consensus was reached. When data were missing or unclear, we contacted study authors by email to provide further details. When data were available in figures but not provided in tables or text, they were extracted from the figures using WebPlotDigitizer (version 4.6–4.8)^1^. If two documents were identified that reported overlapping data from the same study, preference was given to the most complete or recent data, or a peer-reviewed publication over grey literature.

For each article included in the analysis, the following data were recorded:

- *report characteristics:* Covidence identifier, publication URL, first and last authors, journal, publication year, title and figure/table/text from which data were sourced.
- *study characteristics:* study design, NCT identifier, trial phase, trial name (if relevant), and trial location.
- *population characteristics:* mean/median age, and age error and/or age range.
- *intervention characteristics:* developer/manufacturer, drug type and name, immunogen (if relevant), dose, adjuvant (if relevant), and number of doses.

##### Extraction of Efficacy or Effectiveness Data

For each article reporting data on the efficacy or effectiveness of an immunisation product, we extracted the following information: 1) clinical endpoint description used to define RSV-associated illness; 2) the number of participants at risk in the treatment and control groups; 3) the number of RSV-related events in each group; 4) the duration of follow-up; 5) the infecting RSV strain; and 6) the method of diagnosis and strain identification. Where data were reported on both the per-protocol sample and intention-to-treat sample, we extracted the per-protocol data. Efficacy data stratified by age, clinical subgroup, location, gender, ancestry, income level, or weight were excluded. Efficacy data from phase 1 trials (or where there were fewer than 25 participants in the vaccine or control study arms) were not used. Time-to-event data were also excluded. Where we encountered duplicate data reported in several publications, we used the most complete and up-to-date data, preferring data reported in peer-reviewed publications rather than grey literature.

##### Extraction of Immunogenicity Data

For each article reporting immunogenicity data that was selected for inclusion in the analysis, we extracted the following information: 1) aggregate data for neutralising and IgG binding antibodies (anti-RSV or anti-F only) measured at baseline and post-intervention; 2) antibody type measured; 3) number of subjects contributing to each recorded measure; 4) sample type (e.g. serum or nasal fluids); 5) whether data were linear or log-transformed; 6) immunoassay type; 7) RSV strain used in the assay; 8) assay limit of detection; and 9) time between administration of the intervention (or birth) and the measurement. For data that were included in our correlates analysis, we additionally extracted the associated confidence intervals or quartiles of the immunological measurement in both the immunised and control groups at baseline and post-intervention. When both absolute and relative antibody levels were reported, we extracted only the absolute levels. Data from competitive antibody assays were excluded. Immunogenicity data stratified by age, clinical subgroup, location, gender, ancestry, income level, or weight were also excluded. Where we encountered duplicate data reported in several publications, or data from the same samples assayed more than once, we used the most complete and up-to-date data, preferring data reported in peer-reviewed publications rather than grey literature.

#### 1.3. Calculation of Efficacy and 95% Confidence Bounds

We used extracted data on the number of events and subjects at risk in the treatment and control groups for each immunisation product to calculate the efficacy (and associated confidence intervals) against each outcome measure, for each product.

The relative risk was calculated as

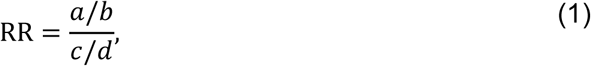

where

*a* = # events in intervention group

*b* = # in intervention group at risk

*c* = # events in reference group

*d* = # in reference group at risk.

The upper and lower confidence intervals of the relative risk on a log-scale, represented by log (RR_CI_), were calculated as

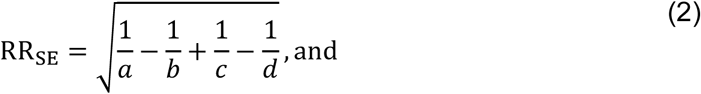

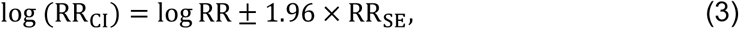

where RR_SE_ represents the standard error of the relative risk on a log-scale. We then calculated efficacy against each outcome (for both the point estimate and 95% confidence interval bounds) as 1 − *RR*.

#### 1.4. Severity Categorisation

The identified studies reporting on vaccine efficacy or effectiveness (hereafter referred to as efficacy) all reported efficacy against different outcome measures. We summarised the outcome measures by the symptoms that were reported to be associated with each outcome.

##### Infant and Children Severity Classification

The outcomes for infants and children were classified into three groups – Symptomatic, Moderate and Severe Disease. Subjects required an RSV-positive PCR test to be classified into any of the three severity categories. Categories from the primary studies were re-categorised into Symptomatic, Moderate or Severe categories when the underlying category from the primary study contained the criteria according to the following requirements:

- Symptomatic:

- at least one reported symptom; <u>or</u>
- seen by a healthcare provider.
- Moderate:

- oxygen saturation <95%; <u>or</u>
- respiratory rate greater than 60, 50, and 40 beats per minute in infants under two months, infants under one year, and children over one year, respectively.
- Severe:

- oxygen saturation < 93%; <u>or</u>
- respiratory rate greater than 70, 60, and 50 beats per minute in infants under two months, under one year, and over one year, respectively.

We note that some studies reported on a mixture of the above classifications (e.g. Dieussaert et. al. reported efficacy against an outcome measure that required <u>either</u> oxygen saturation <93% <u>or</u> respiratory rate greater than 60, 50 and 40 beats per minute in infants under two months, under one year, and children over one year, respectively)^2^. In this situation, we classified the category based on the oxygen saturation criteria rather than the respiratory rate criteria.

##### Older Adult Severity Classification

The outcomes for older adults were classified into three groups based on the number of symptoms a patient required in order to be recorded under each outcome measure – one, two, or three symptoms. Once again, subjects required an RSV-positive PCR test to be classified into any of the three categories.

Two studies (Falsey et. al. 2023 and Falloon et. al. 2017) reported efficacy against two different outcomes requiring two symptoms^3,4^. One required two lower respiratory tract illness (LRTI) symptoms, and the other allowed at least one of the symptoms not to be associated with an LRTI. In this case, we used efficacy data from the outcome measure reporting on two LRTI symptoms.

Two publications (Papi et. al. and Feldman et. al.) reported efficacy against an outcome that required either (i) three symptoms or signs; or (ii) two symptoms or signs, with one of them being a sign^5,6^. We associated this outcome with the three-symptom category.

The resulting dataset, comprised of the efficacy and relative risk (and the corresponding confidence intervals) against the appropriate severity endpoint category for each vaccine product, is henceforth referred to as the *efficacy dataset*.

We calculated the pooled efficacy across immunisation products against each severity endpoint, for each of infants and children, and older adults, using the rma.mv() function with maximum likelihood estimation, from the metafor package version 4.8-0 in R version 4.5.0^7^.

#### 1.5. Estimating the Fold-Rise in Antibody Titre or Concentration

For each immunisation product, we calculated the fold-rise in antibody titre or concentration (either neutralising antibodies or IgG binding antibodies) following immunisation. For the main analysis, we calculated the fold-rise as the difference on a log_10_ scale between the immunised and placebo groups. For maternal vaccine studies in infants, the fold-rise was the difference between immunised and placebo infants at birth. For long-acting monoclonal and older adult vaccine studies, we calculated the fold-rise as the maximum difference in antibody measurement between the immunised and placebo groups following administration, so that

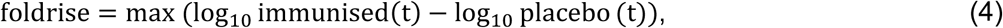

where *t* represents the time in days following birth or administration (Figure S2, panel A). For the studies of older adult vaccines, some studies did not report values for a placebo group; where no placebo group was reported, we used the antibody concentration in the vaccinated group at the time of administration as baseline.

We used the extracted confidence intervals to calculate the variance (on a log_10_ scale) for each antibody measurement. We then calculated the variance of the fold-rise in antibody titres as:

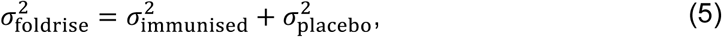

and used the total variance 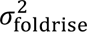 to calculate each 95% confidence interval as

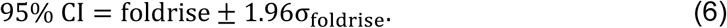

Where neutralising antibody titres were reported against both RSV-A and RSV-B, we observed that the rises in titres were generally similar for both strains. We therefore calculated the total fold-rise in antibody titre as the geometric mean of the fold-rise in neutralising antibody titres against each strain, and the total variance as the mean variance across studies, using the formula

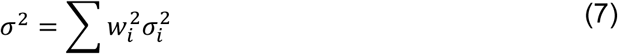

where the weights corresponded to the number of subjects for each antibody measurement, scaled such that ∑ *w*_*i*_ = 1, and calculated the upper and lower 95% confidence bounds using Equation (6) above.

The resulting dataset is henceforth referred to as the *antibody fold-rise dataset*.

We calculated the pooled antibody fold-rise (for each of neutralising titre and IgG concentration) for each immunisation product and for each of infants and children, and older adults, using the rma.mv() function with maximum likelihood estimation, from the metafor package version 4.8-0 in R version 4.5.0^7^.

#### 1.6. Determining the Relationship between Antibodies and Protection from RSV Disease

##### 1.6.1. Combining the Efficacy and Antibody Titre Data

In order to explore the relationship between the rise in antibody levels and protection, we linked each efficacy measurement (in the efficacy dataset) with the corresponding antibody fold-rise (from the antibody fold-rise dataset), for the same immunisation product, formulation, and population group (hereafter referred to only as a vaccine formulation). This produced two combined efficacy-antibody datasets; one for infants and children, and one for older adults.

Where there were multiple immunogenicity studies for a single vaccine formulation, we defined the fold-rise for that vaccine formulation as the weighted average of the fold-rises across all relevant immunogenicity studies, where the weights were given by the number of participants in each study. The upper and lower confidence intervals were similarly calculated using the weighted variances across the studies (see Equation (6)-(7) above).

##### 1.6.1. Model Fitting

In order to quantify the relationship between immunisation-induced antibodies and protection from RSV disease, we fitted a linear mixed effects meta-regression model to the combined efficacy-antibody dataset (restricted to infants and children or older adults only). The model captures the relationship between the fold-rise in neutralising antibody titre (on a log_10_ scale) and the relative risk of disease (on a natural log scale) and optional additional terms to account for the severity of the outcome.

In each model, RR is the relative risk, Rise is the fold-rise in neutralising antibody titre on a log_10_ scale, “:” indicates an interaction term, and “Severity” is a categorical variable describing the severity classification (either Symptomatic, Moderate, or Severe for infants and children, or 1, 2, or 3 symptoms for older adults). A random effect for each study (“Study”) was applied to account for variability in reporting of titres between studies.

We also present each model in mathematical notation, where log(RR)_*s*_ denotes the log of the relative risk of disease from study *s*, β_0_ is the intercept, β_1_ is the fixed effect on the rise, Rise_*s*_ is the antibody fold-rise from study 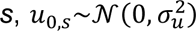 is the random effect on the intercept for study *s*, and ε_*s*_∼*N*(0, σ^2^) is the random error for study *s*.

The “single slope and intercept model” is described by

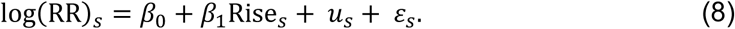

The “varying slope model” is described by

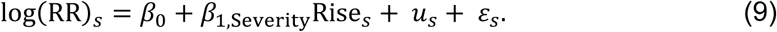

The model was fitted using the rma.mv() function with maximum likelihood estimation, from the metafor package version 4.8-0 in R version 4.5.0^7^.

## 2. Supplementary Sensitivity Analysis

We undertook a range of sensitivity analyses on the analysis of the relationship between the RSV antibody fold-rise and effectiveness in each of the infants and children, and older adults. These models and their associated goodness of fits are shown in Table S6, and the fitted models are depicted in Figures S5 and S6.

### 2.1. Importance of Accounting for Uncertainty in the Antibody Fold-Rise

As an additional sensitivity analysis, we tested the sensitivity of our results to uncertainty in the fold-rise in antibody levels. This was because the statistical model fitting described above using the metafor package accounts for uncertainty in the estimate of conferred protection (y-axis confidence intervals in Figure S4), but does not account for uncertainty in the reported fold-rise in antibody levels (x-axis confidence intervals in Figure S4). We therefore used multiple imputation of the antibody concentration dataset to test the sensitivity of our results to the potential impact of this additional uncertainty. The method is described briefly as follows:

a. Randomly sample from a normal distribution (using the estimated fold-rise as the mean and the 95% confidence interval to calculate the variance) of each estimate of fold-rise in neutralising antibody titre, generating a new dataset for the fold-rise, while leaving efficacy estimates unchanged.
b. Fit the model to this new dataset, using the same procedure as outlined above, and record the set of fitted parameters and the variance-covariance matrix.
c. Repeat steps (a) and (b) 50 times, storing the outputs each time.
d. Combine the estimates of the model parameters and variance using Rubin’s Rules^8^.

This allows us to estimate a new set of fitted parameters 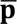 the within-imputation variance *W*, the between-imputation variance *B*, and the total variance *T*, as described in Stadler et. al. 2023^9^ and outlined below in Equations (10)-(13).

The estimate for the new set of fitted parameters is given by

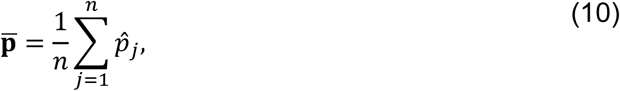

Where *n* is the number of imputed datasets, and 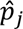 represents the vector of parameters for the *j*th imputed dataset.

The within-imputation variance is given by

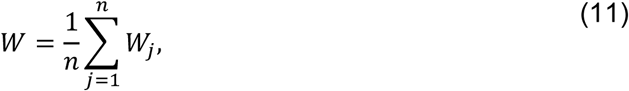

where *W*_*j*_ is the variance-covariance matrix of the estimated parameter set from the *j*th imputed dataset.

The between-imputation variance is given by

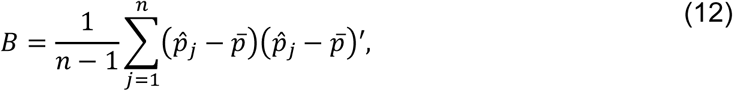

and the total variance is given by

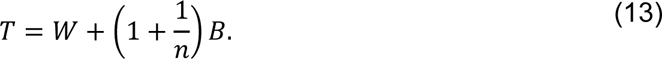

Using the fitted parameters and the total variance, we calculated the 95% confidence intervals for the fitted parameter values, accounting for both x- and y-axis uncertainty.

In order to understand the importance of including the uncertainty in antibody fold-rises in our analysis, we compared the fitted parameters and 95% confidence intervals of selected models with and without multiple imputation. We performed this analysis for the main fitted model for infants and children, and for older adults (“Varying slope model”, Table S6). We found that inclusion of the variation in antibody titres did not change the central estimate of the relationship between antibody fold-rise and efficacy. Antibody fold-rise uncertainty resulted in a negligible increase in the confidence interval around each model parameter (Table S8). We concluded that for both infants and children, and for older adults, most of the variability in the antibody-efficacy relationship in our models arises from uncertainty around the efficacy estimates in our dataset, rather than from uncertainty in the fold-rise in neutralising titre.

### 2.2. Varying the Calculation of Antibody Fold-Rise in the Infants and Children Analysis

As a sensitivity analysis, we considered an alternative measure of the fold-rise in neutralising antibody titre. A limitation of measuring the fold-rise in antibody titre as relative to the placebo group is that we cannot be certain that there are not inherent differences between the immunised and placebo groups within that study. Instead of the fold-rise measured as the difference between the immunised and placebo group, we instead calculated the fold-rise as the maximum increase in antibody titre from the time of administration within the immunised group (Figure S2, panel C).

Note that in infants whose mothers received the maternal vaccination, we do not observe the antibody titre at the time of immunisation. Therefore, for this alternative measure, we calculated the fold-rise in antibodies as the maximum increase in titre from time 0 in the immunised maternal group for the maternal vaccine products. We found that the linear relationship between neutralising titre and the odds of protection against RSV disease was maintained, and that the parameters were similar between the main analysis and fits to this alternative dataset (Table S7). This led us to conclude that our results were not dependent on our choice of how to measure the fold-rise in antibody titre.

## 3. Supplementary Figures

**Figure S1.**
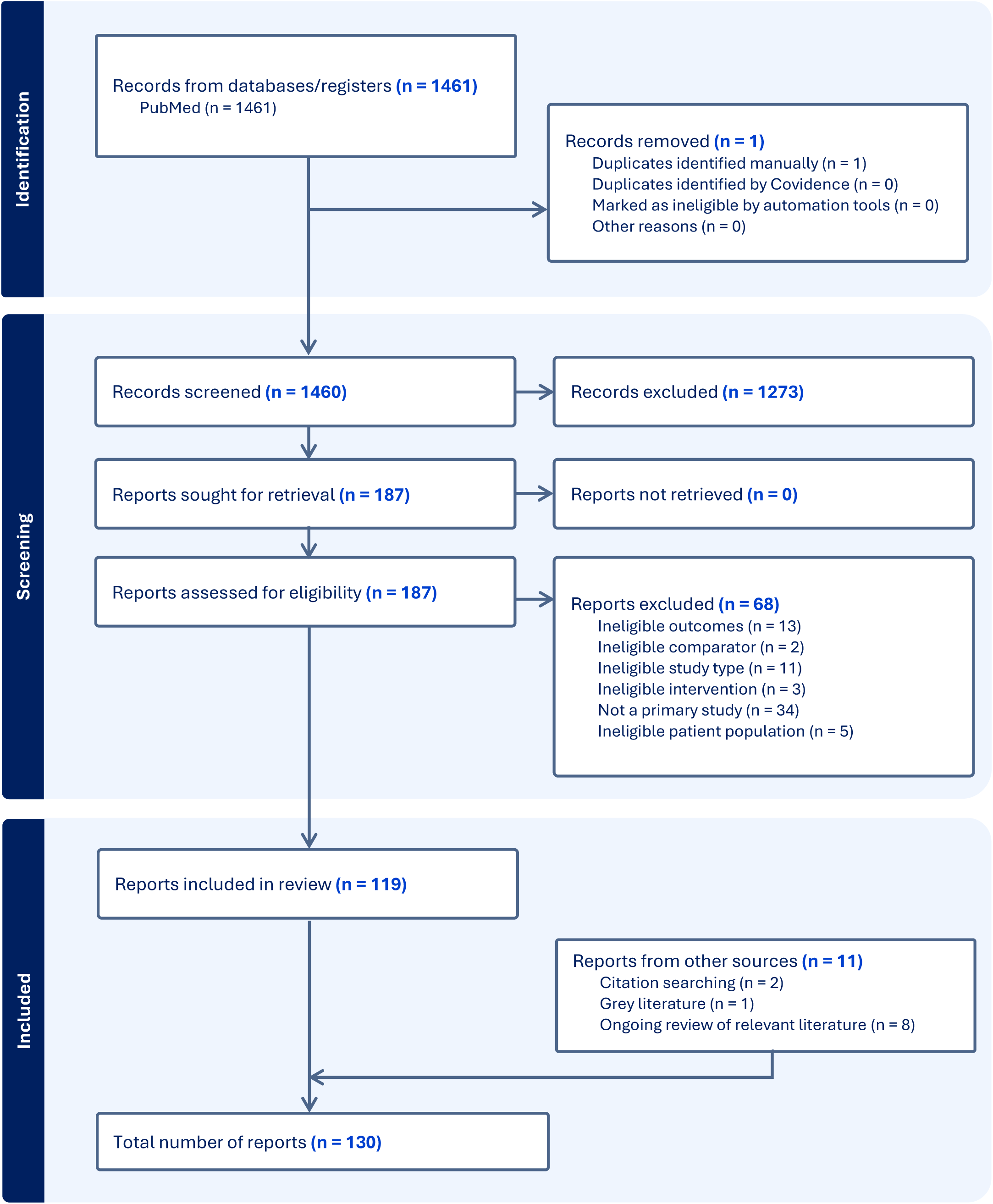
PRISMA flow diagram of report inclusion.

**Figure S2.**
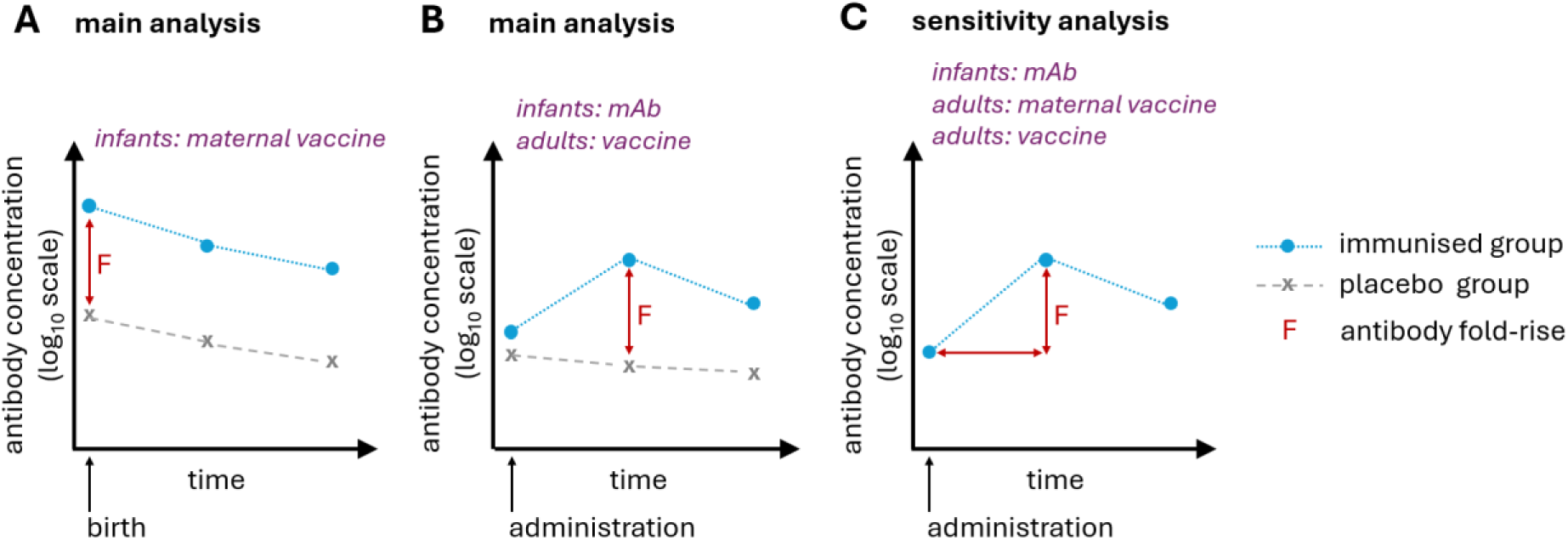
Illustration of how the fold-rise in antibody titre was calculated for different immunisation products and population groups. (A) The fold-rise in antibody titre measured as the difference (on a log-scale) between the antibody titres at birth in infants of mothers who had received the maternal vaccine (immunised group) and who had received a placebo vaccine (placebo group). This was used for infants born to vaccinated mothers. (B) The fold-rise in antibody titre measured as the maximum difference between the antibody titres in the immunised and placebo groups following administration. This was used for vaccinated adults, and infants who received a long-acting monoclonal antibody. The time at which the peak was recorded was, on average, approximately one-month post immunisation. (C) As a sensitivity analysis, we also calculated the fold-rise in antibody titre as the difference between the maximum antibody titre following immunisation and the antibody titre at the time of administration, purely within the immunised group. For this sensitivity analysis, in infants born to vaccinated mothers, this antibody fold-rise was calculated in the (vaccinated) mothers at the time of administration, rather than in the infants at the time of birth.

**Figure S3.**
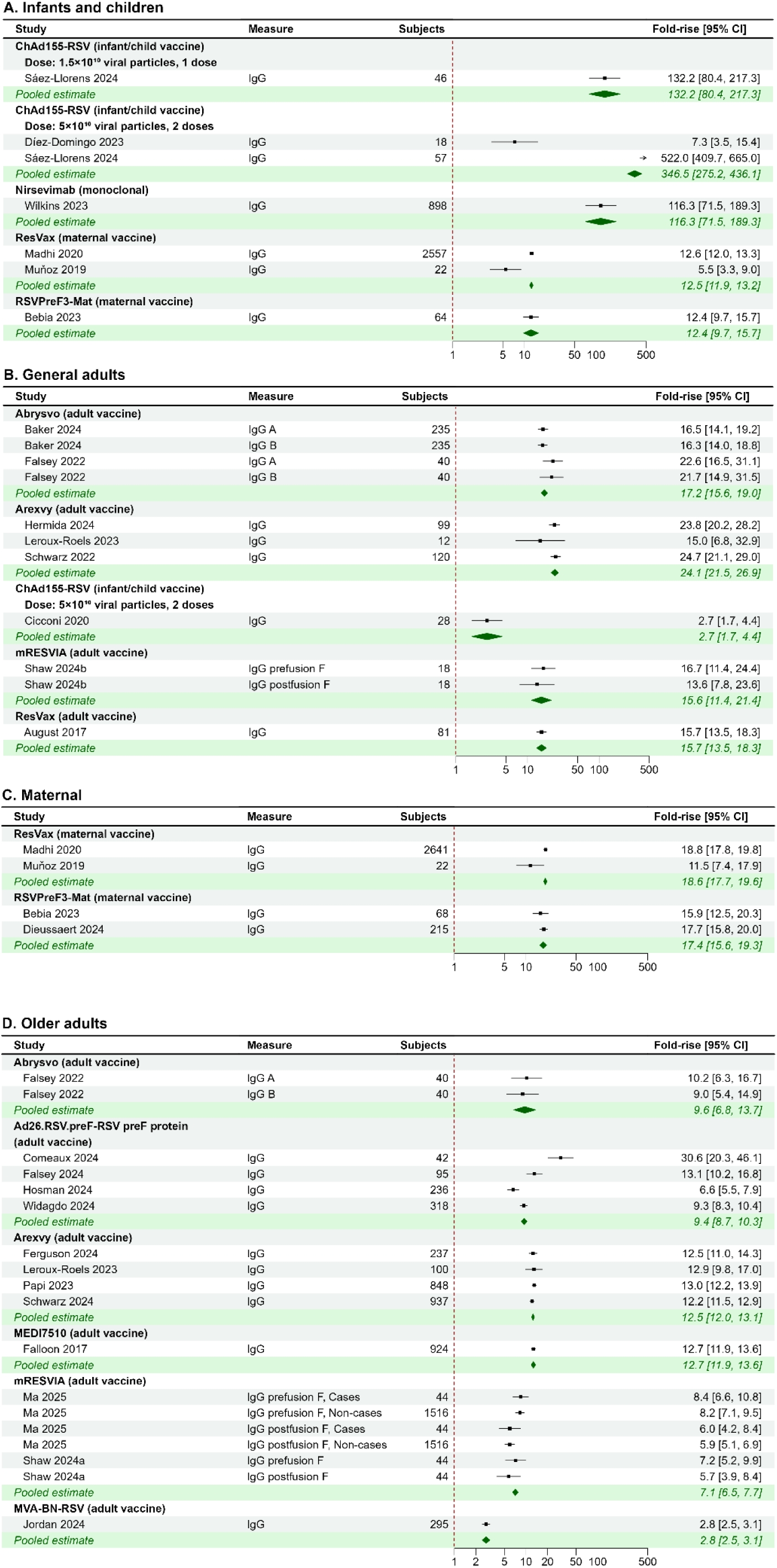
Fold-rise in IgG concentration for different RSV immunisation products in (A) infants and children (6 reports); (B) general adults (8 reports); (C) maternal populations (4 reports); and (D) older adults (13 reports). Individual published reports are grouped by immunisation product, with the pooled estimate for each product shown on the green shaded rows.

**Figure S4.**
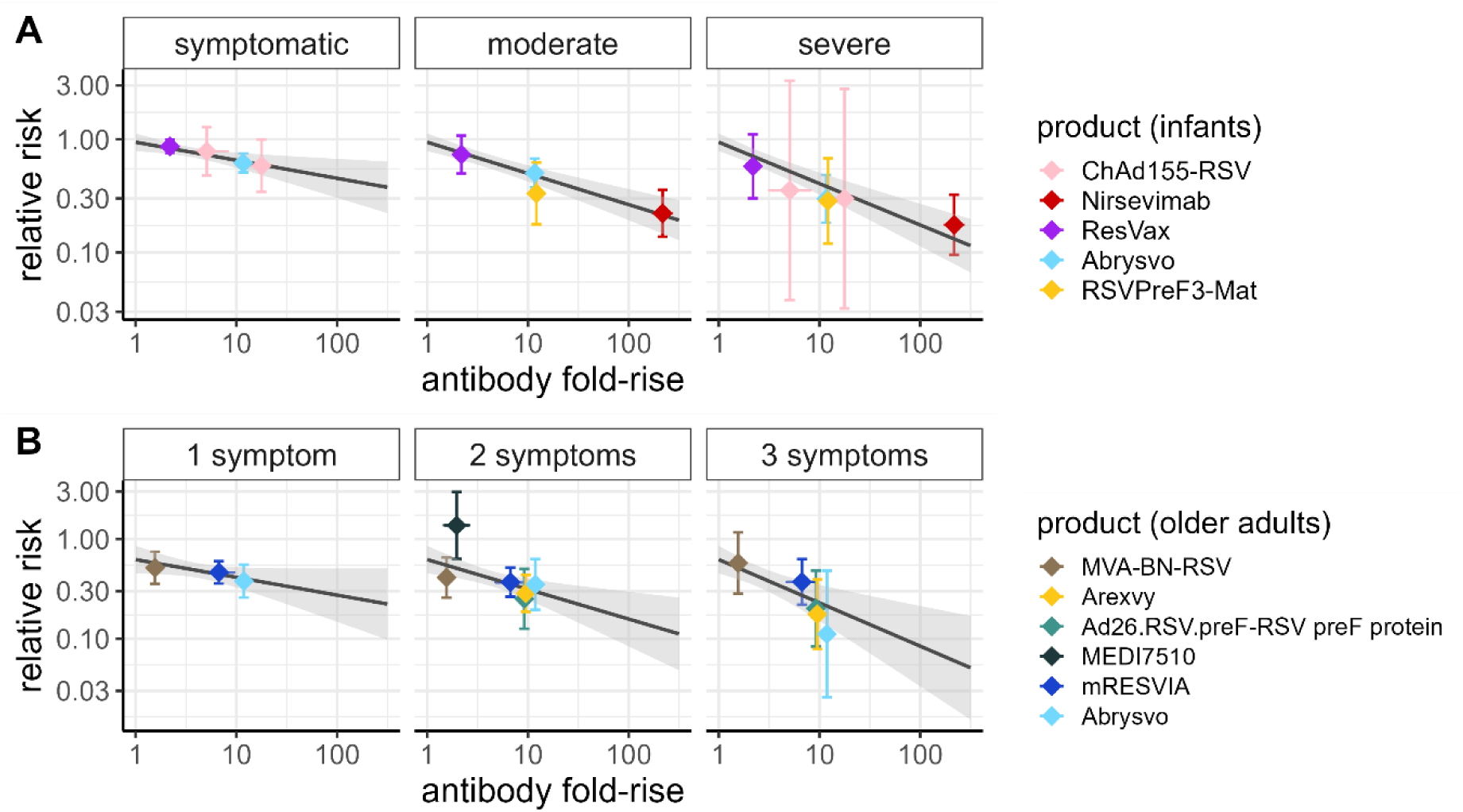
Modelled protection against RSV in infants and children and in older adults using a single intercept and varying slope, corresponding to the efficacy presented in Figure 4 of the main manuscript. Relationship between the fold-rise in neutralising antibody titre and the relative risk of disease in (A) infants and children, against symptomatic, moderate, and severe RSV; and in (B) older adults, against RSV with 1, 2, or 3 symptoms. The immunisation products are shown as coloured diamonds with corresponding confidence intervals. In each panel, the black line represents the fitted statistical model, and grey shaded region represents the 95% confidence interval. For each population group, the fitted model is a model with a single fitted intercept, and a fitted slope for each severity level (Table S6, “Varying slope model”). The models fitted to the data expressed as the efficacy are shown in Figure 4.

**Figure S5.**
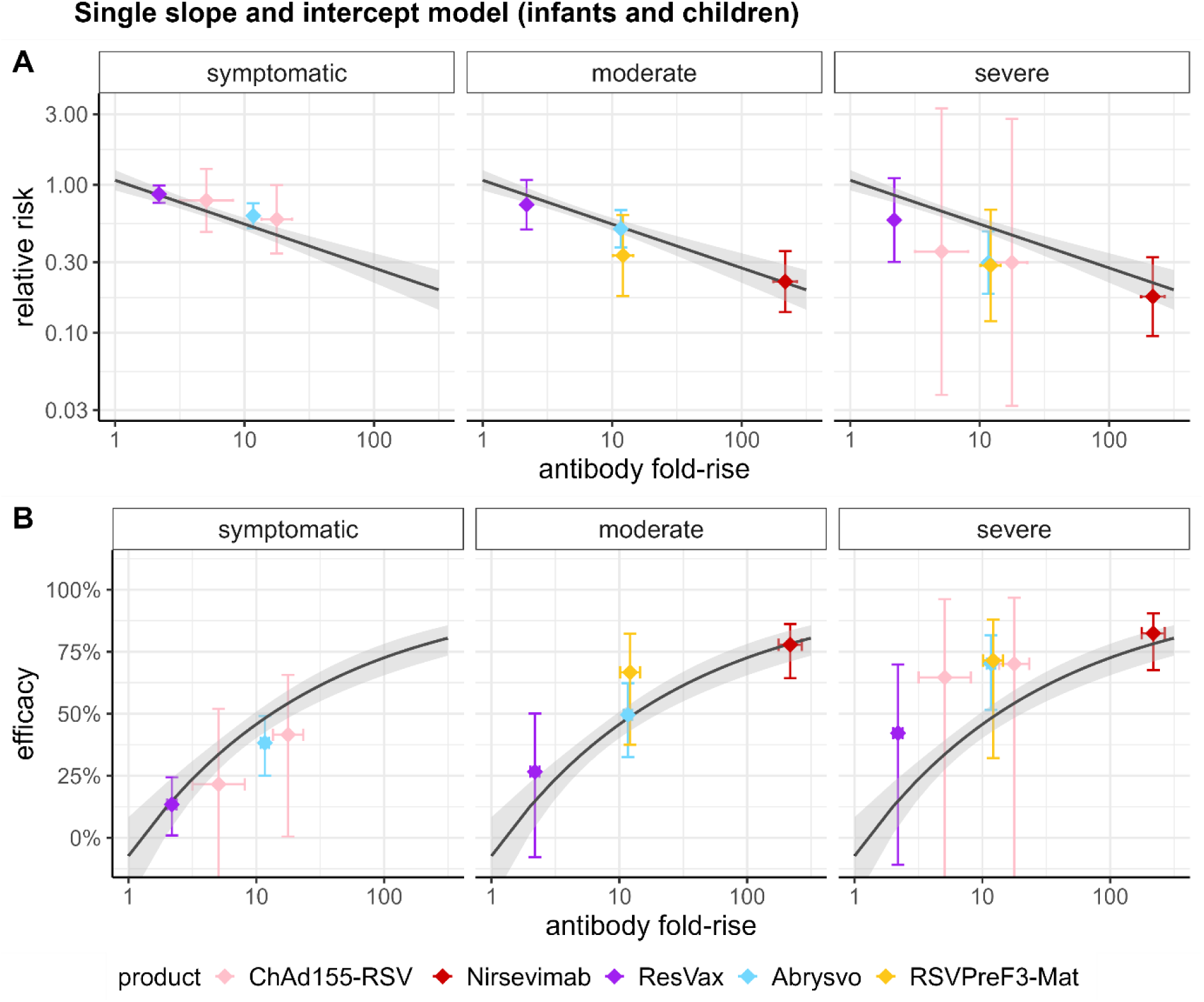
Sensitivity analysis of a “single slope and intercept model” in infants and children. Relationship between the fold-rise in neutralising antibody titre and protection in infants and children, against symptomatic, moderate, and severe RSV, for five immunisation products. Sensitivity analysis illustrating a model with a single fitted slope and intercept (see Table S6). (A) Relationship between antibody fold-rise and the relative risk (*RR*); and (B) Relationship between antibody fold-rise and efficacy, where efficacy is calculated as 1 − *RR*. In each panel, the black line represents the fitted statistical model, and grey shaded region represents the 95% confidence interval.

**Figure S6.**
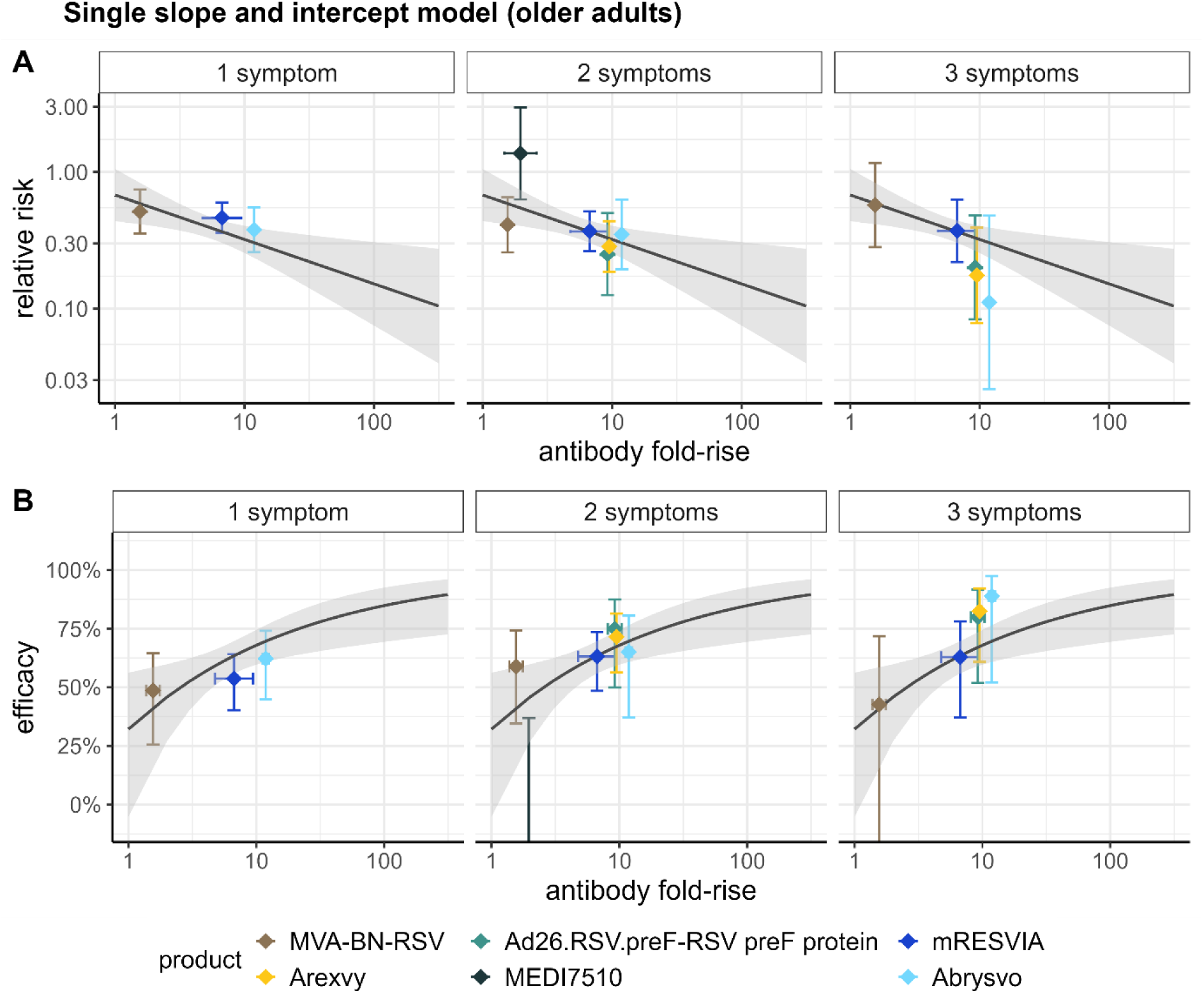
Sensitivity analysis of a “single slope and intercept model” in older adults. Relationship between the fold-rise in neutralising antibody titre and protection in older adults, against three severity outcomes for RSV disease, for six immunisation products. Sensitivity analysis illustrating a model with a single fitted slope and intercept (see Table S6**Table S6**). (A) Relationship between antibody fold-rise and the relative risk (*RR*); and (B) Relationship between antibody fold-rise and efficacy, where efficacy is calculated as 1 − *RR*. Relationship is shown for efficacy against RSV disease with one, two or three symptoms. In each panel, the black line represents the fitted statistical model, and grey shaded region represents the 95% confidence interval.

## 4. Supplementary Tables

**Table S1.** List of scientific drug names for products included in the analysis, alongside drug trade names where applicable. The drug name used in the manuscript is indicated with an asterisk. Throughout the manuscript, the drug scientific name is generally used, unless there is likely to be confusion due to similar scientific names, or if a scientific name is long, in which case the trade name is applied throughout.

| Pharmaceutical developer | Indicated or intended population | Drug scientific name | Drug trade name |
| --- | --- | --- | --- |
| AstraZeneca and Sanofi | Infants | *Nirsevimab | Beyfortus |
| Bavarian Nordic | Older adults | *MVA-BN-RSV |  |
| GSK | Infants and children | *ChAd155-RSV |  |
| GSK | Older adults | RSVPreF3 OA | *Arexvy |
| GSK | Pregnant women | *RSVPreF3-Mat |  |
| Janssen | Older adults | *Ad26.RSV.preF–RSV preF protein |  |
| MedImmune | Older adults | *MEDI7510 |  |
| Moderna | Older adults | mRNA-1345 | *mRESVIA |
| Novavax | Pregnant women | RSV fusion (F) protein nanoparticle vaccine | *ResVax |
| Pfizer | Pregnant women<br>Older adults | RSVpreF | *Abrysvo |

**Table S2.** Reports from the systematic review that reported on efficacy outcomes, and explanations of whether they were included in forest plots and the correlates analysis. Note that the internal report ID (the unique reference number we assigned to each report) corresponds to that in the data and analysis code provided in the Github repository, which will be made available upon publication.

| Reference | Internal report ID | Efficacy assessed in | Efficacy included in forest plot | Efficacy included in correlates analysis |
| --- | --- | --- | --- | --- |
| Díez-Domingo et al 2023<sup>10</sup> | R15 | Children | No: efficacy was reported for aggregated data including multiple dosing regimens | N/A |
| Sáez-Llorens et al 2024<sup>11</sup> | R16 | Infants | Yes | Yes |
| Simões et al 2022<sup>12</sup> | R18 | Infants | No: efficacy was only reported for aggregated data including multiple dosing regimens | N/A |
| Munjal 2023<sup>13</sup> | R21 | Infants | No, efficacy was only reported for aggregated data including multiple dosing regimens | N/A |
| Muller et al 2023<sup>14</sup> | R19 | Infants | Yes | Yes |
| Dagan et al 2024<sup>15</sup> | R20 | Infants | No: data for 1st RSV season is captured in Muller et al 2023<sup>14</sup>; other data is for 2nd RSV season only (excluding 1st season) so not relevant to analysis. | N/A |
| Hammit et al<sup>16</sup> | R80 | Infants | No: data captured in Muller et al 2023<sup>14</sup> | N/A |
| Madhi et al 2020<sup>17</sup> | R22 | Infants | Yes | Yes |
| Kampmann et al 2023<sup>18</sup> | R23 | Infants | Yes: Symptomatic severity outcomes only, as other severity outcomes for the same cohort are captured in Simões et al 2025<sup>19</sup> | Yes |
| Assad et al 2024<sup>20</sup> | R26 | Infants | No: reported on a non-randomised case-control study | N/A |
| Munro et al 2025<sup>21</sup> | R90 | Infants | Yes | Yes |
| Drysdale et al 2023<sup>22</sup> | R25 | Infants | No: data captured in Munro et al 2025<sup>21</sup> | N/A |
| Simões et al 2025<sup>19</sup> | R92 | Infants | Yes: Moderate and Severe outcomes only, as the Symptomatic severity outcome for the same cohort is captured in Kampmann et al 2023<sup>18</sup> | Yes |
| Banooni et al 2025<sup>23</sup> | R93 | Infants | Yes | Yes |
| Dieussaert et al 2024<sup>2</sup> | R24 | Infants | No: data is captured in Banooni et al 2025<sup>23</sup> | N/A |
| Consolati et al 2024<sup>24</sup> | R107 | Infants | No: reported on a non-randomised observational study, and reported risk of RSV bronchiolitis hospitalisation as outcome measure rather than adjusted effectiveness | N/A |
| Estrella-Porter et al 2024<sup>25</sup> | R108 | Infants | No: reported on a non-randomised observational study | N/A |
| Ezpeleta et al 2024<sup>26</sup> | R109 | Infants | No: reported on a non-randomised observational study | N/A |
| López-Lacort et al 2024<sup>27</sup> | R110 | Infants | No: reported on a non-randomised observational study | N/A |
| Moline et al 2024<sup>28</sup> | R111 | Infants | No: reported on a non-randomised observational study | N/A |
| Falsey et al 2023<sup>3</sup> | R45 | Older adults | Yes | Yes |
| Falsey et al 2024<sup>29</sup> | R46 | Older adults | No: data either captured in Falsey et al 2023<sup>3</sup>, or reported over RSV seasons 2 and 3, or 3 RSV seasons, therefore not relevant to analysis | N/A |
| Papi et al 2023<sup>5</sup> | R50 | Older adults | Yes | Yes |
| Feldman et al 2024<sup>6</sup> | R55 | Older adults | No: data captured in Papi et al 2023<sup>5</sup> | N/A |
| Ison et al 2024<sup>30</sup> | R58 | Older adults | No: same cohort as in Papi et al 2023<sup>5</sup> but over different follow-up periods | N/A |
| Falloon et al 2017<sup>4</sup> | R62 | Older adults | Yes | Yes |
| Jordan et al 2024<sup>31</sup> | R88 | Older adults | Yes | Yes |
| Ma et al 2025<sup>32</sup> | R89 | Older adults | Yes | Yes |
| Wilson et al 2023<sup>33</sup> | R42 | Older adults | No: data captured in Ma et al 2025<sup>32</sup> | N/A |
| Walsh et al 2024a<sup>34</sup> | R91 | Older adults | Yes | Yes |
| Walsh et al 2025<sup>35</sup> | R94 | Older adults | No: data captured in Walsh et al 2024<sup>34</sup> | N/A |
| Walsh et al 2023<sup>36</sup> | R41 | Older adults | No: data captured in Walsh et al 2024<sup>34</sup> | N/A |
| Curran et al 2024<sup>37</sup> | R102 | Older adults | No: data captured in Papi et al 2023<sup>5</sup> | N/A |
| Schmoele-Thoma et al 2022<sup>38</sup> | R56 | General adults | No: human challenge study | N/A |
| Sadoff et al 2022<sup>39</sup> | R60 | General adults | No: human challenge study | N/A |
| Jordan et al 2023<sup>40</sup> | R61 | General adults | No: human challenge study | N/A |

**Table S3.** Reports from the systematic review that reported on antibody titres, and explanations of whether they were included in forest plots and correlates analysis. Note that the internal report ID (the unique reference number we assigned to each report) corresponds to that in the data and analysis code provided in the Github repository, which will be made available upon publication.

| Reference | Internal report ID | Antibody titres assessed in | Antibody titres included in forest plots | Antibody titres included in correlates analysis |
| --- | --- | --- | --- | --- |
| Bebia et al 2023<sup>41</sup> | R02 | Maternal and infants | Yes | Infants: yes<br>Maternal: in sensitivity analysis |
| Buchholz et al 2018<sup>42</sup> | R03 | Infants/children | No: Did not match formulation used in an identified efficacy study | N/A |
| Karron et al 2020<sup>43</sup> | R04 | Infants/children | No: Did not match formulation used in an identified efficacy study | N/A |
| Cunningham et al 2019<sup>44</sup> | R05 | Infants | No: Did not match formulation used in an identified efficacy study | N/A |
| Karron et al 2015<sup>45</sup> | R06 | General adults and children | No: Did not match formulation used in an identified efficacy study | N/A |
| Karron et al 2024<sup>46</sup> | R07 | Infants/children | No: Did not match formulation used in an identified efficacy study | N/A |
| Malkin et al 2013<sup>47</sup> | R08 | Infants | No: Did not match formulation used in an identified efficacy study | N/A |
| McFarland et al 2018<sup>48</sup> | R09 | Infants | No: Did not match formulation used in an identified efficacy study | N/A |
| McFarland et al 2020a<sup>49</sup> | R10 | Infants | No: Did not match formulation used in an identified efficacy study | N/A |
| McFarland et al 2020b<sup>50</sup> | R11 | Infants | No: Did not match formulation used in an identified efficacy study | N/A |
| Stuart et al 2023<sup>51</sup> | R12 | General adults and children | No: Did not match formulation used in an identified efficacy study | N/A |
| Cunningham et al 2022<sup>52</sup> | R13 | Infants/children | No: Did not match formulation used in an identified efficacy study | N/A |
| Wilkins et al 2023<sup>53</sup> | R14 | Infants | Yes | Yes |
| Díez-Domingo et al 2023<sup>10</sup> | R15 | Children | Yes | Yes |
| Sáez-Llorens et al 2024<sup>11</sup> | R16 | Infants | Yes | Yes |
| Muñoz et al 2019<sup>54</sup> | R17 | Maternal and infants | Yes | Infants: yes<br>Maternal: in sensitivity analysis |
| Simões et al 2022<sup>12</sup> | R18 | Maternal and infants | Infants: Yes<br>Maternal: No, as maternal data are reported at ~2 months post-administration, and are captured in Munjal 2023<sup>13</sup> at 1 month post-administration | Yes |
| Munjal 2023<sup>13</sup> | R21 | Maternal and infants | Maternal: Yes<br>Infants: No, as data captured in Simões et al 2022<sup>12</sup> | No efficacy data in maternal populations; but used in infants sensitivity analysis |
| Madhi et al 2020<sup>17</sup> | R22 | Maternal and infants | Yes | Infants: Yes<br>Maternal: In sensitivity analysis |
| Dieussaert et al 2024<sup>2</sup> | R24 | Maternal and infants | Infants: No, as data captured in Banooni et al 2025<sup>23</sup><br>Maternal: Yes (IgG only, as neutralising titre captured in Banooni et al 2025<sup>23</sup>) | N/A |
| Abarca et al 2020<sup>55</sup> | R27 | General adults | No: Did not match formulation used in an identified efficacy study | N/A |
| Ascough et al 2019<sup>56</sup> | R30 | General adults | No: Did not match formulation used in an identified efficacy study | N/A |
| Athan et al 2024<sup>57</sup> | R31 | Older adults | Yes | Yes |
| August et al 2018<sup>58</sup> | R32 | General adults | Yes | No relevant efficacy data available in general adults |
| Baber et al 2022<sup>59</sup> | R33 | Older adults | No: Did not match formulation used in an identified efficacy study | N/A |
| Baker et al 2024<sup>60</sup> | R34 | General adults | Yes | No relevant efficacy data available in general adults |
| Cicconi et al 2020<sup>61</sup> | R39 | General adults | Yes | No relevant efficacy data available in general adults |
| Falsey et al 2022<sup>62</sup> | R40 | General and older adults | Yes | General adults: No relevant efficacy data available in general adults<br>Older adults: Yes |
| Falsey et al 2023<sup>3</sup> | R45 | Older adults | Yes | Yes |
| Falsey et al 2024<sup>63</sup> | R46 | Older adults | Yes | Yes |
| Shaw et al 2024a<sup>64</sup> | R47 | Older adults | Yes | Yes |
| Shaw et al 2024b<sup>65</sup> | R48 | General adults | Yes | No relevant efficacy data available in general adults |
| Schwarz et al 2024<sup>66</sup> | R49 | Older adults | Yes | Yes |
| Papi et al 2023<sup>5</sup> | R50 | Older adults | Yes | Yes |
| Leroux-Roels et al 2024<sup>67</sup> | R51 | Older adults | No: Did not match formulation used in an identified efficacy study | N/A |
| Leroux-Roels et al 2023a<sup>68</sup> | R52 | General and older adults | Yes | General adults: No relevant efficacy data available in general adults<br>Older adults: Yes |
| Ferguson et al 2024<sup>69</sup> | R53 | Older adults | Yes<sup>a</sup> | No: Did not report neutralising antibody titres. |
| Kotb et al 2023<sup>70</sup> | R54 | Older adults | No: Did not match formulation used in an identified efficacy study | N/A |
| Feldman et al 2024<sup>6</sup> | R55 | Older adults | No: Data captured in Papi et al 2023<sup>5</sup> | N/A |
| Schmoele-Thoma et al 2022<sup>38</sup> | R56 | General adults | No: Human challenge study | N/A |
| Walsh et al 2022<sup>71</sup> | R57 | General adults | No: Data captured in Falsey et al 2022<sup>62</sup> | N/A |
| Walsh et al 2024b<sup>72</sup> | R59 | General and older adults | Did not match formulation used in an identified efficacy study | N/A |
| Sadoff et al 2022<sup>39</sup> | R60 | General adults | No: Human challenge study | N/A |
| Jordan et al 2023<sup>40</sup> | R61 | General adults | No: Human challenge study | N/A |
| Falloon et al 2017a<sup>4</sup> | R62 | Older adults | Yes | Yes |
| Falloon et al 2017b<sup>73</sup> | R63 | Older adults | No: Did not match formulation used in an identified efficacy study | N/A |
| Hermida et al 2024<sup>74</sup> | R64 | General adults | Yes | No relevant efficacy data available in general adults |
| Beran et al 2018<sup>75</sup> | R65 | General adults | No: Did not match formulation used in an identified efficacy study | N/A |
| Schwarz et al 2019<sup>76</sup> | R66 | General adults | No: Did not match formulation used in an identified efficacy study | N/A |
| Comeaux et al 2024<sup>77</sup> | R67 | Older adults | Yes | Yes |
| Jordan et al 2021<sup>78</sup> | R69 | Older adults | No: Efficacy data for test formulation limited to human challenge study | N/A |
| Robbie et al 2013<sup>79</sup> | R70 | General adults | No: Did not match formulation used in an identified efficacy study | N/A |
| Leroux-Roels et al 2019<sup>80</sup> | R71 | General adults | No: Did not match formulation used in an identified efficacy study | N/A |
| Schwarz et al 2022<sup>81</sup> | R72 | General adults | Yes | No relevant efficacy data available in general adults |
| Hosman et al 2024<sup>82</sup> | R73 | Older adults | Yes | Yes |
| Sadoff et al 2021<sup>83</sup> | R74 | Older adults | No: Efficacy data for test vaccine limited to human challenge study | N/A |
| Williams et al 2020<sup>84</sup> | R75 | Older adults | No: Efficacy data for test vaccine limited to human challenge study | N/A |
| Langley et al 2017<sup>85</sup> | R77 | General adults | No: Efficacy data for test formulation limited to human challenge study | N/A |
| Chandler et al 2024<sup>86</sup> | R78 | Older adults | Yes | Yes |
| Peterson et al 2022<sup>87</sup> | R79 | General adults | No: No corresponding efficacy data identified by review for test regimen (co-administered with another vaccine) | N/A |
| Fries et al 2017<sup>88</sup> | R81 | Older adults | No: Efficacy data for test formulation limited to human challenge study | N/A |
| Glenn et al 2016<sup>89</sup> | R82 | General adults | No: Efficacy data for test formulation limited to human challenge study | N/A |
| Glenn et al 2013<sup>90</sup> | R83 | General adults | No: Efficacy data for test formulation limited to human challenge study | N/A |

<sup>a</sup> Note this was a randomised study without a control arm, reporting on binding antibody titres only.

| Reference | Internal report ID | Antibody titres assessed in | Antibody titres included in forest plots | Antibody titres included in correlates analysis |
| --- | --- | --- | --- | --- |
| Ruckwardt et al 2021<sup>91</sup> | R85 | General adults | No: Efficacy data for test formulation limited to human challenge study | N/A |
| Spearman et al 2023<sup>92</sup> | R86 | General and older adults | No: Efficacy data for test formulation limited to human challenge study | N/A |
| Griffin et al 2017<sup>93</sup> | R87 | General adults | No: Efficacy data for test formulation limited to human challenge study | N/A |
| Jordan et al 2024<sup>31</sup> | R88 | Older adults | Yes | Yes |
| Ma et al 2025<sup>32</sup> | R89 | Older adults | Yes | No: Stratified by cases and non-cases |
| Simões et al 2025<sup>19</sup> | R92 | Maternal and infants | Yes: (Infants only)<br>Maternal: No, as data reported only at administration and delivery | Yes |
| Banooni et al 2025<sup>23</sup> | R93 | Maternal and infants | Yes | Infants: Yes<br>Maternal: In sensitivity analysis |
| Walsh et al 2025<sup>35</sup> | R94 | Older adults | Yes | Yes |
| Ferguson et al 2024<sup>69</sup> | R53 | Older adults | Yes | No: Only reported binding antibody titres |
| Weinberg et al 2019<sup>94</sup> | R68 | Older adults | No: F-specific IgA data not relevant to analysis, and IgG data assumed to be captured in Falloon et al (R62) | N/A |
| Widagdo et al 2023<sup>95</sup> | R76 | Older adults | Yes | No: Only reported binding antibody titres |
| Leroux-Roels et al 2023b<sup>96</sup> | R84 | General adults | No: Did not match formulation used in an identified efficacy study | N/A |
| Falloon et al 2016<sup>97</sup> | R95 | Older adults | No: Did not match formulation used in an identified efficacy study | N/A |
| Chang et al 2022<sup>98</sup> | R96 | General and older adults | Older adults: Yes<br>General adults: Data for a product for which no efficacy data available | Older adults: Yes |
| Pacheco et al 2023<sup>99</sup> | R97 | General adults | No: Did not match formulation used in an identified efficacy study | N/A |
| Green et al 2015<sup>100</sup> | R98 | General adults | No: Did not match formulation used in an identified efficacy study | N/A |
| Green et al 2019<sup>101</sup> | R99 | General and older adults | No: Did not match formulation used in an identified efficacy study | N/A |
| Sacconnay et al 2023<sup>102</sup> | R100 | Older adults | No: Data already captured in Leroux-Roels et al 2023a<sup>68</sup> | N/A |
| Crank et al 2019<sup>103</sup> | R101 | General adults | No: Did not match formulation used in an identified efficacy study | N/A |
| Gerretsen et al 2018<sup>104</sup> | R103 | General adults | No: Did not match formulation used in an identified efficacy study | N/A |
| Aliprantis et al 2021a<sup>105</sup> | R104 | General and older adults | No: Did not match formulation used in an identified efficacy study | N/A |
| Aliprantis et al 2021b<sup>106</sup> | R105 | General adults | No: Did not match formulation used in an identified efficacy study | N/A |
| Orito et al 2021<sup>107</sup> | R106 | General adults | No: Did not match formulation used in an identified efficacy study | N/A |
| Mukhamedova 2021<sup>108</sup> | R115 | General adults | No: Did not match formulation used in an identified efficacy study | N/A |
| Samy et al 2020<sup>109</sup> | R117 | General adults | No: Did not match formulation used in an identified efficacy study | N/A |
| Verdijk et al 2020<sup>110</sup> | R118 | General adults | No: Did not match formulation used in an identified efficacy study | N/A |

**Table S4.** Reports included in the correlates of protection analysis in infants and children.

| Manufacturer | Drug and type | Dose | Adjuvant | Efficacy studies (Internal report ID) | Immunogenicity studies (Internal report ID) |
| --- | --- | --- | --- | --- | --- |
| AstraZeneca & Sanofi | Nirsevimab monoclonal antibody | 50mg or 100mg | None | Muller et al 2023<sup>14</sup> (R19)<br>Munro et al 2025<sup>21</sup> (R90) | Wilkins et al 2023<sup>53</sup> (R14) |
| GSK | RSVPreF3-Mat maternal vaccine | 120µg | None | Banooni et al 2025<sup>23</sup> (R93) | Bebia et al 2023<sup>41</sup> (R02)<br>Banooni et al 2025<sup>23</sup> (R93) |
| GSK | ChAd155-RSV infants and children vaccine | 1.5×10<sup>10</sup> viral particles (1 dose);<br>5×10<sup>10</sup> viral particles, (2 doses) | None | Sáez-Llorens et al 2024<sup>11</sup> (R16) | Sáez-Llorens et al 2024<sup>11</sup> (R16)<br>Díez-Domingo et al 2023<sup>10</sup> (R15) |
| Novavax | ResVax maternal vaccine | 120µg | 0.4mg Al | Madhi et al 2020<sup>17</sup> (R22) | Muñoz et al 2019<sup>54</sup> (R17)<br>Madhi et al 2020<sup>17</sup> (R22) |
| Pfizer | Abrysvo maternal vaccine | 120µg | None | Kampmann et al 2023<sup>18</sup> (R23)<br>Simões et al 2025<sup>19</sup> (R92) | Simões et al 2022<sup>12</sup> (R18)<br>Simões et al 2025<sup>19</sup> (R92)<br>Munjal 2023<sup>13A</sup> (R21) |

<sup>A</sup>Included only for the sensitivity analysis of the antibody fold-rise measure in infants; see Supplementary Methods S2.3

**Table S5.** Reports included in the correlates of protection analysis in older adults.

| Manufacturer | Drug and type | Dose | Adjuvant | Efficacy studies (Internal report ID) | Immunogenicity studies (Internal report ID) |
| --- | --- | --- | --- | --- | --- |
| Bavarian Nordic | MVA-BN-RSV vaccine | 3x10<sup>8</sup> infectious units | None | Jordan et al 2024<sup>31</sup> (R88) | Jordan et al 2024<sup>31</sup> (R88) |
| GSK | Arexvy vaccine | 120µg | AS01E | Papi et al 2023<sup>5</sup> (R50) | Schwarz et al 2024<sup>66</sup> (R49)<br>Papi et al 2023<sup>5</sup> (R50)<br>Leroux-Roels et al 2023a<sup>68</sup> (R52)<br>Chandler et al 2024<sup>86</sup> (R78) |
| Janssen | Ad26.RSV.preF-RSV preF protein combination vaccine | 1x10<sup>11</sup> viral particles per 150µg protein | None | Falsey et al 2023<sup>3</sup> (R45) | Falsey et al 2023<sup>3</sup> (R45)<br>Falsey et al 2024<sup>29</sup> (R46)<br>Comeaux et al 2024<sup>77</sup> (R67)<br>Hosman et al 2024<sup>82</sup> (R73) |
| MedImmune | MEDI7510 vaccine | 120µg | 5µg GLA | Falloon et al 2017<sup>4</sup> (R62) | Falloon et al 2017<sup>4</sup> (R62)<br>Chang et al 2022<sup>98</sup> (R96) |
| Moderna | mRESVIA vaccine | 50µg | None | Ma et al 2025<sup>32</sup> (R89) | Shaw et al 2024a<sup>64</sup> (R47) |
| Pfizer | Abrysvo vaccine | 120µg | None | Walsh et al 2024a<sup>34</sup> (R91) | Athan et al 2024<sup>57</sup> (R31)<br>Falsey et al 2022<sup>62</sup> (R40)<br>Walsh et al 2025<sup>35</sup> (R94) |

**Table S6.** Details of the models fitted to the immunogenicity-efficacy dataset in infants and children, and in older adults. Full details of the model formulations are in S1.6.2. AIC: Akaike Information Criterion.

| Population | Model name | Model description | Model formulation | Fitted parameters<br>Point estimate (95% CI) | AIC | Results shown in |
| --- | --- | --- | --- | --- | --- | --- |
| Infants and children | Single slope and intercept model | Slope: Fitted (same for all severities)<br>Intercept: Fitted (same for all severities) | Equation (8) | Slope: -0.68 (-0.85, -0.51)<br>Intercept: 0.07 (-0.09, 0.23) | 11.7 | Figure S5 |
| Infants and children | Varying slope model | Slope: Fitted (varies by severity)<br>Intercept: Fitted (same for all severities) | Equation (9) | Slope, Symptomatic: -0.37 (-0.63, -0.1)<br>Slope, Moderate: -0.63 (-0.83, -0.44)<br>Slope, Severe: -0.84 (-1.08, -0.6)<br>Intercept: -0.06 (-0.24, 0.12) | 4.4 | Figure 4;<br>Figure S4 |
| Older adults | Single slope and intercept model | Slope: Fitted (same for all severities)<br>Intercept: Fitted (same for all severities) | Equation (8) | Slope: -0.75 (-1.29, -0.21)<br>Intercept: -0.39 (-0.83, 0.05) | 14.9 | Figure S6 |
| Older adults | Varying slope model | Slope: Fitted (varies by severity)<br>Intercept: Fitted (same for all severities) | Equation (9) | Slope, 1 Symptom: -0.41 (-0.84, 0.02)<br>Slope, 2 Symptoms: -0.69 (-1.12, -0.25)<br>Slope, 3 Symptoms: -1.00 (-1.55, -0.45)<br>Intercept: -0.48 (-0.79, -0.16) | 12.7 | Figure 4;<br>Figure S4 |

**Table S7:** Fitted parameters for the modelled relationship between antibody fold-rise and efficacy in infants for two measures of the fold-rise in infants and children. The fitted parameters are for the “Varying slope model” in infants and children (Table S6). The main analysis calculates the fold-rise in neutralising antibody titre following birth or immunisation as relative to the placebo group, whereas in a sensitivity analysis, we calculate the fold-rise as relative to the value in the immunised group at the time of vaccine administration, i.e., time 0 (see Supplementary Methods S1.5 and S2.3).

| Model parameter | Estimated value (95% confidence interval) |  |
| --- | --- | --- |
|  | Data with fold-rise relative to placebo (main analysis) | Data with fold-rise relative to time 0 (sensitivity analysis) |
| Slope, Classification: Symptomatic | -0.37 (-0.63, -0.1) | -0.38 (-0.63, -0.13) |
| Slope, Classification: Moderate | -0.63 (-0.83, -0.44) | -0.64 (-0.84, -0.44) |
| Slope, Classification: Severe | -0.84 (-1.08, -0.6) | -0.88 (-1.13, -0.63) |
| Intercept | -0.06 (-0.24, 0.12) | -0.02 (-0.21, 0.17) |

**Table S8.** Sensitivity analysis capturing uncertainty in both the fold-rise in neutralising titre and protection in each of infants and children, and older adults. This table shows a comparison of the estimated slopes for the mixed effects meta-regression model in the infants and children population group, and in the older adults population group, where only uncertainty in protection (y-axis uncertainty) is accounted for, and the estimated values for the model parameters where multiple imputation is applied to account for the additional uncertainty in antibody titre (x-axis uncertainty). For each population group model, the fitted parameters are for the “Varying slope model” (Table S6).

| Model and parameter | Estimated value (95% confidence interval) |  |
| --- | --- | --- |
|  | Meta-regression model with uncertainty in protection | Meta-regression model with uncertainty in both protection and antibody fold-rise |
| **Infants and children** |  |  |
| Slope, Classification: Symptomatic | -0.37 (-0.63, -0.1) | -0.37 (-0.63, -0.1) |
| Slope, Classification: Moderate | -0.63 (-0.83, -0.44) | -0.64 (-0.83, -0.44) |
| Slope, Classification: Severe | -0.84 (-1.08, -0.6) | -0.84 (-1.08, -0.6) |
| Intercept | -0.06 (-0.24, 0.12) | -0.06 (-0.24, 0.12) |
| **Older adults** |  |  |
| Slope, Classification: 1 Symptom | -0.41 (-0.84, 0.02) | -0.4 (-0.83, 0.03) |
| Slope, Classification: 2 Symptoms | -0.69 (-1.12, -0.25) | -0.68 (-1.12, -0.25) |
| Slope, Classification: 3 Symptoms | -1 (-1.55, -0.45) | -0.99 (-1.55, -0.43) |
| Intercept | -0.48 (-0.79, -0.16) | -0.48 (-0.8, -0.17) |

**Table S9:** Summary of calculated efficacy and protection results for each of infants and children, and older adults. These results correspond to those described in the text and figures from the main manuscript. Results are presented along with the 95% confidence intervals (in brackets). Model: “Varying slope” model; RR: Relative Risk

|  | Pooled efficacy across products identified in systematic review | Reduction in RR of disease for each 10-fold rise in neutralising antibody titres |
| --- | --- | --- |
| **Estimate derived from** | Meta-analysis | Model fit |
| **Manuscript location** | Figure 2 and Results text | Results text |
| **Infants and children** |  |  |
| Symptomatic | 23.3% [14.7, 31.0] | 30.7% [9.7, 46.9] |
| Moderate | 53.6% [43.5, 62.0] | 47.0% [35.5, 56.4] |
| Severe | 69.8% [59.2, 77.7] | 56.9% [45.2, 66.2] |
| **Older adults** |  |  |
| 1 symptom | 54.7% [45.6, 62.3] | 33.7% [-2.0, 56.9] |
| 2 symptoms | 62.7% [54.5, 69.3] | 49.7% [22.3, 67.4] |
| 3 symptoms | 69.2% [57.0, 78.0] | 63.2% [36.0, 78.8] |

**Table S10:** Summary of immunisation induced rises in neutralising antibody titres in adult cohorts of different ages, for three adult vaccine products (Abrysvo, Arexvy and mRESVIA). Values correspond to those reported in Table 3 in the main text and show pooled estimates and 95% confidence intervals (in brackets). The final column shows the fold-change in the immunisation induced rise in antibody titres in younger versus older adults. A positive fold-change difference indicates that the immunisation induced rise in neutralising titres is higher in younger adults than in older adults.

| Product | Age range | Combined estimate for immunisation-induced rise in neutralising antibody titres [95% CI] | Fold-change in immunisation-induced rise in neutralising antibody titres in younger versus older adults [95% CI] |
| --- | --- | --- | --- |
| Abrysvo | 18-49 years<sup>60,62</sup> | 14.5 [13.0, 16.3] | 1.2 [1.1, 1.4] |
|  | >60 years<sup>35,57,62</sup> | 11.8 [11.2, 12.5] |  |
| Arexvy | 18-45 years<sup>68,74,81</sup> | 17.0 [14.7, 19.7] | 1.7 [1.5, 2.0] |
|  | >60 years<sup>5,66,86,96</sup> | 9.8 [9.5, 10.1] |  |
| mRESVIA | 18-49 years<sup>65</sup> | 13.0 [8.1, 20.9] | 1.8 [1.1, 2.9] |
|  | >60 years<sup>32,64</sup> | 7.3 [6.5, 8.2] |  |

## Notes

### Competing Interest Statement

The authors have declared no competing interest.

### Author Declarations

This study used only publicly available data, extracted from the published reports identified in the systematic review.

